# Therapist effects in real-world rehabilitation outcomes: a cohort study of the nationwide GLA:D osteoarthritis management program in Denmark

**DOI:** 10.64898/2026.04.20.26351120

**Authors:** Phillips Edomwonyi Obasohan, Joe Palmer, David Alderson, Dahai Yu, Dorte T. Grønne, Ewa M. Roos, Søren T. Skou, George M. Peat

## Abstract

**Objective:** Unlike several other fields of healthcare, little is known about the size of ‘therapist effects’ on patient outcomes following rehabilitation for musculoskeletal conditions. We aimed to estimate the proportion of variance in patient outcomes from a structured rehabilitation program explained by therapist effects.

**Methods:** For our observational cohort study we accessed data from the national multicentre Good Life with osteoArthritis in Denmark (GLA:D) osteoarthritis management program. Analyses included 23,021 consecutive eligible adults with hip or knee osteoarthritis (mean (SD) age 65.0 (9.8) years, 71% female) treated by 657 therapists between October 2014 and February 2019. The primary outcome was ≥30% reduction in pain intensity on 0-100 VAS at 3 months. Therapist effects were estimated as the variance partition coefficient (intra-class correlation coefficient (ICC)) from two-level random intercept logistic regression models before and after adjusting for patient-level case-mix factors and therapist-level characteristics (number of patients treated, days since therapist certification). Analyses were repeated for a range of secondary outcomes using multiply imputed data and complete-case analysis.

**Results:** 52% of patients reported a ≥30% reduction in pain intensity on 0-100 VAS at 3 months. In the null model the ICC was 0.007 (95%CI: 0.005, 0.009), which changed little after adjusting for patient- and therapist-level covariates. Upper confidence limits for ICC estimates across all secondary outcomes in multiply imputed and complete case analyses were less than 0.03.

**Conclusions:** In a nationally implemented osteoarthritis management program delivered by trained healthcare professionals, therapist effects made a minimal contribution to variation in patient outcomes.

**KEY MESSAGES:** *What is already known on this topic:* ‘Therapist effects’ - defined as the effect of a given therapist on patient outcomes as compared to another therapist - have been observed in several fields of healthcare and have important consequences for selection, training, and service improvement. In musculoskeletal rehabilitation five previous studies suggest that 1-12% of variation in patient-reported outcomes may be attributable to therapist effects, but these estimates were based on relatively small datasets resulting in substantial uncertainty.

*What this study adds:* Our cohort study analysed registry data from 2014-2019 on 23,021 patients and 647 trained therapists from the nationally implemented GLA:D structured osteoarthritis management program in Denmark. We found that therapist effects accounted for less than 3% of total variation in patient-reported pain and quality of life outcomes 3 months after beginning the program

*How this study might affect research, practice, or policy:* Our findings suggest that contextual factors that relate to therapist effects – therapist characteristics or therapist-patient interaction and alliance - make a minimal contribution to variation in patient outcomes from this structured, group-based rehabilitation intervention. Any contextual effects must be attributable to alternative sources, e.g. patient expectations, intervention setting.

## INTRODUCTION

The range, difficulty, and variability of behaviours required of health professionals are recognised as key mechanisms through which many complex healthcare interventions produce patient benefit.[1] Therefore, studies that set out to estimate the magnitude of “therapist effects” - defined as the effect of a given therapist on patient outcomes as compared to another therapist[2] - have the potential to drive important new insights and lead to improvements in care: Johns et al. (2019)[3] list four potential ways that studies of therapist effects contribute to knowledge and care: they temper an over-emphasis on ‘brands’ of treatment; they identify more or less effective therapists with implications for matching patients, as well as selection, training, and revalidation; they contribute to mechanistic understanding; and they generate research questions designed to reduce unwarranted variability in service provision and outcomes.

In psychotherapy, surgery, and medicine, where there is a relatively long history of such studies, there is strong evidence that therapist effects contribute to patient outcomes, although estimates vary substantially. Whilst extreme estimates suggest that between 0 and 47% of the variation in patient outcomes may be attributable to therapist effects, the more plausible range consistently observed is 3-10%.[3–7] Factors such as the volume of procedures undertaken by individual surgeons[8] or psychotherapists’ interpersonal skills[9] have been proposed as potential determinants.

A comparable body of evidence is largely absent for musculoskeletal rehabilitative interventions. We found only five studies, conducted over the last 15 years, which have estimated therapist effects in this field (**Table 1**).[10–14]

**Table 1:** Previously published estimates of therapist effects in musculoskeletal rehabilitation.

| Author, year | Study Design | Setting | Participants | Therapists/<br>practitioners | Intervention(s) | Outcomes | Model | ICC/<br>VPC | Comments |
| --- | --- | --- | --- | --- | --- | --- | --- | --- | --- |
| Lewis et al.,<br>2010 | Secondary<br>analysis of 3<br>RCTs | Physiotherapy<br>services (UK) | 350 adults with<br>neck pain | 38 PTs | Multiple (advice<br>and exercise;<br>manual therapy;<br>pulsed<br>shortwave<br>diathermy) | Self-reported<br>functional limitation<br>(NPDQ) and<br>psychological health<br>(SF-12 MH) at 6w, 6m | Multi-level (patients<br>nested in therapists)<br>linear and logistic<br>regression models<br>applied to each<br>outcome and<br>endpoint | 0.008-0.071 | Greater therapist<br>effect<br>hypothesised<br>when treatment<br>was standardised |
|  |  | Primary care<br>(NL) | 314 adults with low<br>back pain | 54 GPs | Psychosocial<br>intervention;<br>usual care | Self-reported<br>functional limitation<br>(RMDQ) and<br>psychological health<br>(FABQ) at 6w, 3m,<br>12m |  | 0.012 –<br>0.08 |  |
|  |  | Primary care<br>(UK) | 402 adults with low<br>back pain | 6 PTs | Brief<br>psychologically-<br>informed pain<br>management;<br>manual therapy | Self-reported<br>functional limitation<br>(RMDQ) and<br>psychological health<br>(TSK) at 3m, 12m |  | 0.027 –<br>0.051 |  |
| Simon et al.,<br>2013 | Observational<br>cohort | Outpatient<br>physical<br>therapy (US) | 258 adults with<br>neck pain or low<br>back pain | 5 PTs | Usual care:<br>multiple<br>treatments<br>(mainly manual<br>therapy) | Self-reported pain<br>intensity (VAS) and<br>function (CCFOI) at<br>discharge | Multi-level (patients<br>nested in therapists)<br>linear regression<br>models | ≤0.035 | No model<br>convergence for<br>pain |
| Buining et al.,<br>2015 | Observational<br>cohort | Primary care<br>(NL) | 393 adults with<br>non-reversible NCD<br>(CVD, RA,<br>orthopaedic,<br>neurological;<br>including 180 with<br>OA) | 39 PTs | Usual care | Self-reported<br>symptom severity (0-10 NRS) at end of<br>treatment | Multi-level (patients<br>nested in therapists)<br>linear regression<br>model | 0.076 | Outcome<br>positively<br>associated with<br>low PT<br>neuroticism |
| Kooijman et al., 2020 | Observational cohort | Primary care (NL) | 1013 adults with rotator cuff pain | 46 PTs | Usual care | Change in self-reported severity of complaint (0-10 NRS) between start and end of treatment | Multi-level (patients nested in therapists?) linear regression model | 0.12 | Outcome positively associated with PT extraversion |
| Nudelman et al., 2025 | Observational cohort | Outpatient physical therapy (ISR) | 1043 adults with low back pain | 68 PTs | Usual care | Self-reported functional status (FS) score (0-100 LCAT) and residual (risk-adjusted) FS score at discharge | Multi-level (patients nested in therapists) linear regression model | 0.10 (FS score)<br>0.04 (residual FS score) | Outcome negatively associated with PT biomedically-oriented attitude but clinical relevance questionable |
| **CCFOI** CareConnections functional outcomes index; **CVD** Cardiovascular disease; **FABQ** Fear-Avoidance Beliefs Questionnaire; **FS** Functional Scale; **GP** General practice/practitioner; **ICC** Intraclass correlation coefficient; **ISR** Israel; **LCAT** Lumbar Computerized Adaptive Test; **NCD** Non-communicable disease; **NL** Netherlands; **NPDQ** Northwick Park Disability Questionnaire; **NRS** Numerical rating scale; **OA** Osteoarthritis; **PT** Physical therapist/physiotherapist; **RA** Rheumatoid arthritis; **RCT** Randomised controlled trial; **RMDQ** Roland-Morris Disability Questionnaire; **SF-12 MH** Short-form 12 Mental Health; **TSK** Tampa Scale of Kinesiophobia; **VAS** Visual Analog Scale; **VPC** Variance Partition Component; **UK** United Kingdom |  |  |  |  |  |  |  |  |  |

None met the suggested requirement for multi-level models of therapist effects that studies include outcomes from at least 100 healthcare professionals treating at least 10 patients each.[3] One reason for the apparent paucity of evidence in musculoskeletal rehabilitation has been a lack of available large-scale registry data that combine the collection of standardised patient outcome measures and unique identifiers for therapists. The national rollout of structured osteoarthritis management programs with accompanying registry data created the opportunity to examine therapist effects in this field. The nationwide Good Life with osteoArthritis in Denmark (GLA:D®) program is an 8-week structured, group-based, physiotherapist-led osteoarthritis management program for people with hip or knee osteoarthritis comprising 2-3 patient education sessions and 12 clinician-supervised exercise therapy sessions. Since beginning in 2013, tens of thousands of patients have taken part in programs delivered by trained physiotherapists across Denmark, mainly in primary care centres and municipal settings.[15]

Given the unique opportunity afforded by the de-identified data from the GLA:D registry, our aim was to estimate ‘real world’ therapist effects on patient-reported outcomes from a structured, supervised group-based musculoskeletal rehabilitation intervention.

## METHODS

### Data source

The Danish national, electronic GLA:D® registry houses data on participant characteristics and outcomes collected at baseline, three months, and 12 months via a combination of patient-reported, therapist-reported, and objective measures, and the routine collection of standard outcomes is an integral component of the GLA:D® program. The GLA:D® registry was approved by the Danish Data Protection Agency. According to the Danish Data Protection Act, patient consent is not required as personal data was processed exclusively for research and statistical purposes. Separate ethics approval was not needed for the current analysis.

### Population

For the current analysis all consecutive participants with hip or knee OA enrolled on the GLA:D® program in Denmark between 9 October 2014 and 28 February 2019 were potentially eligible. These dates represent a period before COVID-19 during which the outcome measures, exposures, and covariates of interest in this analysis were included in the data-collection instruments. Participants who had not returned a patient-reported questionnaire at baseline or who did not have a completed therapist ID were excluded from our analyses. We included only participants with a baseline pain intensity score ≥40 out of 100, a common eligibility criterion for clinical trials in osteoarthritis.[16,17] For participants taking the program more than once, only the first (index) attendance was included in the analysis.

### Outcomes

The primary outcome of interest was clinically important pain reduction at 3 months, defined as ≥30% reduction in pain intensity (0-100 VAS) between baseline and 3 months.[17,18] Secondary outcomes were ≥50% reduction in pain intensity (0-100 VAS), pain intensity score (0-100 VAS) at 3 months, HOOS/KOOS Quality of Life subscale score (0-100) at 3 months, EQ5D Health Utility Score (5L Danish values, -0.757 – 1.000) at 3 months, and EQ5D Health-Related Quality of Life VAS (0-100) at 3 months. Pain and quality of life (hip/knee-related and general health-related) represent two of the five domains for core outcome sets in hip and knee osteoarthritis trials (the others being physical function, patient global assessment, and adverse events),[19,20] and are among highly-rated optional recommended domains for evaluating osteoarthritis management programs[21]

### Therapists

All certified healthcare practitioners delivering the GLA:D intervention complete 2-day standardised training. The majority are physiotherapists. Analyses were restricted to therapists who had treated at least 10 patients.[3] Detailed information on therapists is not routinely collected. However, we used cumulative number of patients treated and number of days since completing GLA:D training as indicators of therapist experience.

### Patient-level covariates for case-mix adjustment

Differences in case-mix may explain apparent therapist effects. For case-mix adjustment in our analyses we identified the following potential patient-level determinants of pain outcome from previous literature[22] and previous analyses of GLA:D data[23–26]: patient age, sex, born in Denmark, Danish citizen, month and year of entry to GLA:D program, most affected joint (hip/knee), body mass index (kg/m^2^), duration of symptoms, walking speed time during 40 metre walk test at baseline (m/s), number of chairs stands completed in 30 seconds at baseline, number of painful body areas at baseline (0-56), UCLA physical activity score at baseline (1-10), EQ-5D VAS at baseline (0-100), EQ-5D Health Utility score (5L Danish values - 0.757 - 1.000), KOOS/HOOS Quality of Life score at baseline (0-100), Arthritis Self-Efficacy Scale Pain and Other subscale scores (10-100), SF-12 Physical and Mental Health Component Scores (0-100), pain intensity (0-100) at baseline.

### Statistical analysis

The rate and pattern of missing data were evaluated using the *naniar* package in R. Rates of missing data ranged from 0% to 30% (HOOS/KOOS QOL score at 3 months) due mainly to loss to follow-up (27%) and removal/later inclusion of symptom duration and SF-12 baseline between 2014 and 2019 (i.e. missing by design) (**Supplementary data**). Our primary analysis was of multiply imputed data, judging that complete case analysis or simple imputation were insufficient due to the risk of bias.[27] Imputation models were based on the assumption of data missing at random (MAR). Given the concentration of missingness in a few variables, we analysed patterns and probabilities of missing data to identify predictors for missing values.[28,29] The imputation model included all variables in the final model, including outcome and auxiliary variables (secondary outcomes).[30] Missing data were imputed using multiple imputation with chained equations using the *mice* package in R to create 40 imputed datasets, to equal or exceed the fraction of missing data[31] and using 10 iterations each. To achieve model convergence, continuous predictors were standardised and centred.

For binary outcomes, we fitted two-level (patients at level 1 nested within therapists at level 2) random intercept logistic regression models (**Table 2**). Three models were fitted. Model 0representes the ‘null’ model with no covariates. Model 1 includes the patient-level (level-1) covariates. Model 2 is the full model consisting of Model 1 and the therapist-level (level-2) covariates. In each model we reported the Variance Partition Coefficient (VPC) estimated by the intra-class correlation coefficient (ICC) using the latent variable method, together with model fit statistics (Akaike Information Criterion (AIC), Bayesian Information Criterion (BIC), and area under the Receiver Operator Curve (AUC)). For continuous secondary outcomes, random-intercept linear regression models were fitted in the same sequence and using AIC and BIC to evaluate model fit.

**Table 2:** Structure of multilevel model (2 levels, primary outcome)

| Level | Sub-index | Variables |
| --- | --- | --- |
| Therapist (657) | $i=1,...,m$ | Number of patients treated on GLA:D, time since date of GLA:D certification |
| Patient (23,021) | $j=1,...,n_i$ | Outcome: $\geq 30\%$ reduction in pain VAS at 3 months<br>Age, sex, Month of entry to GLA:D program, Year of entry to GLA:D program, Most affected joint (hip, knee), BMI, Duration of symptoms, 40m walk test at baseline, Number of chair stands during 30sec at baseline, Number of painful body areas (0-56) at baseline, UCLA - Physical activity score (1-10) at baseline, KOOS/HOOS quality of life score (0-100) at baseline, ASES Other score (10-100) at baseline, ASES Pain score (10-100) at baseline, EQ-VAS (0-100) at baseline, EQ-5D Health Utility score (-0.757-1.000) at baseline, SF-12 Physical Component Score (0-100) at baseline, SF-12 Mental Component Score (0-100) at baseline, pain intensity (0-100 VAS) at baseline |

Sample population Patients attending GLA:D OAMP in Denmark Oct 2014-Feb 2019
Level 2: Treating therapist ( $i=m$ members)
Level 1: Patient ( $j=n_i$ members per unit $i$ )
``` graph TD; SP[Sample population Patients attending GLA:D OAMP in Denmark Oct 2014-Feb 2019] --> T1[Therapist_ID 1]; SP --> Dots1[....]; SP --> Tm[Therapist_ID m]; T1 --> P11[Patient_ID 1.1]; T1 --> Dots2[....]; T1 --> P1n1[Patient_ID 1.n1]; Tm --> Pm1[Patient_ID m.1]; Tm --> Dots3[....]; Tm --> Pmn1[Patient_ID m.n1]; ```

To visually display variation between therapists in patient outcomes we produced funnel plots of the case-mix-adjusted outcomes (therapist-specific observed - expected outcome + overall mean outcome) by number of patients seen per therapist. 95% and 99.8% confidence limits were corrected for an overdispersion factor (phi), calculated using observed variance/expected variance.[32,33]

We compared the above models and outputs from multiply imputed datasets with the same analyses conducted on a complete case dataset of therapists with at least 10 patients, each with fully observed data on all predictors and outcomes.

As a final step, we conducted an exploratory descriptive comparison of patient outcomes for selected anonymised therapists who were identified as potential outliers from the above funnel plots.

Analyses were conducted in R4.4.0. A list of packages used in R is provided in **Supplementary data.**

### Patient and public involvement

Patients and members of the public were not involved in the conceptualisation, design, analysis, interpretation, or dissemination of this study.

## RESULTS

**Table 3** provides the descriptive characteristics of 657 therapists and 23,021 patients included in the current analysis. The median number of patients per therapist was 26 (IQR 16, 44) and therapists were a median of 640 days post-certification (IQR 305, 1073). Patients had a mean (SD) age of 65.0 (9.8) years, 71% were female, and they had typically attended higher (post-secondary) education (59%), and most were either retired (53%) or employed/student (29%). The knee joint was the primary complaint in 74% of patients.

**Table 3.** Descriptive characteristics of patients (n=23,021) and therapists (n=657)

|  | Valid N | Miss % | N | % |
| --- | --- | --- | --- | --- |
| **Therapists** |  |  |  |  |
| Number of patients treated: median (IQR) | 657 | 0 | 36 (16, 72) |  |
| Days since therapist certification: median (IQR) | 657 | 0 | 645 (313, 1077) |  |
| Patients |  |  |  |  |
| Age (years): mean (SD) | 23,021 | 0 | 65.0 (9.8) |  |
| Female sex | 23,021 | 0 | 17,177 | 74.4 |
| Educational level | 22,994 | <1 |  |  |
| Primary |  |  | 4,745 | 20.6 |
| Secondary |  |  | 2,651 | 11.5 |
| Higher – short |  |  | 4,705 | 20.5 |
| Higher – mid |  |  | 8,819 | 38.4 |
| Higher – long |  |  | 2,073 | 9.0 |
| Employment status | 23,020 | <1 |  |  |
| Currently employed/student |  |  | 6,737 | 29.3 |
| Full-time sick leave |  |  | 664 | 2.9 |
| Part-time sick leave |  |  | 691 | 3.0 |
| Retired |  |  | 12,248 | 53.2 |
| Unemployed |  |  | 558 | 2.4 |
| Self-imposed early retirement |  |  | 1,226 | 5.3 |
| Early retirement due to workability issues |  |  | 896 | 3.9 |
| Born in Denmark | 23,009 | <1 | 22,081 | 96.0 |
| Danish citizen | 23,009 | <1 | 37,768 | 98.3 |
| Body mass index (kg/m²): mean (SD) | 22,956 | <1 | 29.0 (5.5) |  |
| Symptom duration (months): median (IQR) | 20,408 | 11 | 18 (6, 48) |  |
| 40m walk test at baseline (m/sec): mean (SD) | 21,792 | 5 | 1.4 (0.3) |  |
| Chair test at baseline (no.): mean (SD) | 22,158 | 4 | 11.4 (3.8) |  |
| No. of pain areas at baseline: mean (SD) | 22,851 | <1 | 4.0 (3.3) |  |
| UCLA Activity score (0-10): mean (SD) | 23,004 | <1 | 5.5 (1.8) |  |
| Primary complaint | 23,019 | <1 |  |  |
| Hip |  |  | 5,928 | 26.0 |
| Knee |  |  | 17,091 | 74.0 |
| Month of enrolment on GLA:D | 23,021 | 0 |  |  |
| January |  |  | 2,618 | 11.4 |
| February |  |  | 1,747 | 7.6 |
| March |  |  | 1,907 | 8.3 |
| April |  |  | 1,985 | 8.6 |
| May |  |  | 1,742 | 7.6 |
| June |  |  | 1,313 | 5.7 |
| July |  |  | 1,012 | 4.4 |
| August |  |  | 2,895 | 12.6 |
| September |  |  | 2,221 | 9.6 |
| October |  |  | 2,727 | 11.8 |
| November |  |  | 1,602 | 7.0 |
| December |  |  | 1,252 | 5.4 |
| Year of enrollment on GLA:D | 23,021 | 0 |  |  |
| 2014 |  |  | 1,214 | 5.3 |
| 2015 |  |  | 3,342 | 14.5 |
| 2016 |  |  | 5,147 | 22.4 |
| 2017 |  |  | 4,981 | 21.6 |
| 2018 |  |  | 4,480 | 19.5 |
| 2019 |  |  | 3,857 | 16.8 |
| Type of organisation | 23,021 | 0 |  |  |
| Public clinic |  |  | 4,042 | 17.6 |
| Private clinic |  |  | 18,974 | 82.4 |
| Private hospital |  |  | 5 | <1 |
| Arthritis Self-Efficacy Scale: Pain at baseline (10-100): mean (SD) | 23,002 | <1 | 61.9 (20.0) |  |
| Arthritis Self-Efficacy Scale: Other at baseline (10-100): mean (SD) | 22,998 | <1 | 66.6 (18.3) |  |
| SF-12 Physical Component Summary at baseline (0-100): mean (SD) | 19,076 | 17 | 35.3 (8.2) |  |
| SF-12 Mental Component Summary at baseline (0-100): mean (SD) | 19,076 | 17 | 51.0 (10.1) |  |
| Pain intensity at baseline (0-100): mean (SD) | 23,021 | 0 | 61.3 (14.5) |  |
| Pain intensity at 3 months (0-100): mean (SD) | 16,746 | 27 | 41.4 (22.6) |  |
| Experienced ≥30% reduction in pain at 3 months | 16,746 | 27 | 8,759 | 52.3 |
| Experienced ≥50% reduction in pain at 3 months | 16,746 | 27 | 5,750 | 34.3 |
| HOOS/KOOS QOL at baseline (0-100): mean (SD) | 22,215 | 4 | 40.9 (14.3) |  |
| HOOS/KOOS QOL at 3 months (0-100): mean (SD) | 16,221 | 30 | 47.8 (16.6) |  |
| EQ5D VAS at baseline (0-100): mean (SD) | 23,003 | <1 | 64.8 (19.0) |  |
| EQ5D VAS at 3 months (0-100): mean (SD) | 16,748 | 27 | 69.7 (19.2) |  |
| EQ5D utility score at baseline (-0.757-1.000): mean (SD) | 22,996 | <1 | 0.710 (0.216) |  |
| EQ5D utility score at 3 months (-0.757-1.000): mean (SD) | 16,743 | 27 | 0.780 (0.204) |  |
| Figures represent N (%) unless otherwise stated<br>Miss % = rate of missing data for each variable<br>**GLA:D** Good Life with osteoArthritis in Denmark; **EQ5D** EuroQoL-5 dimensions; **HOOS** Hip Osteoarthritis Outcome Score; **IQR** Interquartile range; **KOOS** Knee Osteoarthritis Outcome Score; **QOL** Quality of Life; **SD** Standard deviation; **UCLA** University of California, Los Angeles; **VAS** Visual Analogue Score |  |  |  |  |

Pain intensity (0-100 VAS) at baseline was typically moderate to severe (mean 61.3, SD 14.5). Of 16,746 participants with pain intensity scores available at baseline and 3 months, 8,759 (53%) had experienced a 30% or greater reduction in pain intensity at 3 months; 5,750 (34%) experienced a 50% or greater reduction in pain intensity.

In the variance partition coefficient model for the primary outcome in multiply imputed data, the estimated intra-class correlation coefficient was 0.007 (0.005, 0.009) (**Table 4**).

**Table 4.** Random intercept logistic regression models for ≥30% pain reduction at 3 months: multiply imputed data.

|  | Model 0 | Model 1 | Model 2 |
| --- | --- | --- | --- |
|  | 'Null' model | Patient-level adjustment model | 'Full model' |
|  | N=23,021 | N=23,021 | N=23,021 |
|  |  | aOR (95%CI) | aOR (95%CI) |
| **Therapist-level variables** |  |  |  |
| Number of patients treated |  |  | 0.97 (0.93, 1.02) |
| Days since therapist certification |  |  | 0.99 (0.94, 1.04) |
| **Patient-level variables** |  |  |  |
| Age |  | 0.94 (0.90, 0.97) | 0.94 (0.90, 0.97) |
| Female sex |  | 1.11 (1.03, 1.19) | 1.11 (1.03, 1.20) |
| Born in Denmark |  | 1.05 (0.85, 1.29) | 1.04 (0.85, 1.29) |
| Danish citizen |  | 1.05 (0.76, 1.46) | 1.05 (0.76, 1.46) |
| Month of enrollment on GLA:D (ref: January) |  |  |  |
| February |  | 0.96 (0.83, 1.11) | 0.96 (0.83, 1.11) |
| March |  | 1.12 (0.98, 1.29) | 1.13 (0.98, 1.30) |
| April |  | 1.08 (0.95, 1.24) | 1.08 (0.95, 1.24) |
| May |  | 1.16 (1.00, 1.34) | 1.16 (1.01, 1.35) |
| June |  | 1.07 (0.91, 1.26) | 1.08 (0.92, 1.27) |
| July |  | 1.05 (0.89, 1.24) | 1.06 (0.89, 1.25) |
| August |  | 1.09 (0.95, 1.24) | 1.09 (0.96, 1.25) |
| September |  | 1.07 (0.93, 1.22) | 1.07 (0.94, 1.23) |
| October |  | 1.13 (0.99, 1.28) | 1.14 (1.00, 1.29) |
| November |  | 1.15 (0.99, 1.33) | 1.16 (1.00, 1.34) |
| December |  | 0.94 (0.80, 1.10) | 0.95 (0.81, 1.12) |
| Year of enrollment on GLA:D (ref: 2014) |  |  |  |
| 2015 |  | 0.94 (0.81, 1.10) | 0.95 (0.82, 1.11) |
| 2016 |  | 0.88 (0.76, 1.02) | 0.90 (0.78, 1.05) |
| 2017 |  | 1.00 (0.86, 1.16) | 1.04 (0.89, 1.21) |
| 2018 |  | 1.05 (0.90, 1.23) | 1.11 (0.94, 1.31) |
| 2019 |  | 0.97 (0.83, 1.14) | 1.03 (0.86, 1.24) |
| Most affected joint is hip (ref: knee) |  | 0.82 (0.77, 0.88) | 0.82 (0.77, 0.88) |
| BMI (kg/m<sup>2</sup>) |  | 0.95 (0.92, 0.99) | 0.95 (0.92, 0.99) |
| Duration of symptoms (months) |  | 0.91 (0.88, 0.94) | 0.91 (0.88, 0.94) |
| 40m walk test at baseline |  | 1.10 (1.05, 1.14) | 1.10 (1.05, 1.15) |
| No. of chair stands during 30sec at baseline |  | 1.05 (1.01, 1.09) | 1.05 (1.01, 1.09) |
| No. of painful body areas (0-56) at baseline |  | 0.92 (0.88, 0.95) | 0.92 (0.88, 0.95) |
| UCLA - Physical activity (1-10) at baseline |  | 0.99 (0.96, 1.02) | 0.99 (0.96, 1.02) |
| KOOS/HOOS QOL (0-100) at baseline |  | 1.11 (1.06, 1.15) | 1.11 (1.06, 1.15) |
| ASES: Other (10-100) at baseline |  | 0.97 (0.92, 1.03) | 0.97 (0.92, 1.03) |
| ASES: Pain (10-100) at baseline |  | 1.26 (1.20, 1.33) | 1.26 (1.20, 1.33) |
| EQ-VAS (0-100) at baseline |  | 1.08 (1.04, 1.13) | 1.08 (1.04, 1.13) |
| EQ-5D Health Utility score (-0.757-1) at baseline |  | 1.20 (1.15, 1.26) | 1.20 (1.15, 1.26) |
| SF-12 PCS (0-100) at baseline |  | 1.00 (0.96, 1.05) | 1.00 (0.96, 1.05) |
| SF-12 MCS (0-100) at baseline |  | 0.94 (0.90, 0.99) | 0.94 (0.90, 0.99) |
| Pain intensity (0-100) at baseline |  | 1.63 (1.57, 1.69) | 1.63 (1.57, 1.69) |
| VPC (ICC) | 0.007<br>(0.005, 0.009) | 0.008<br>(0.005, 0.011) | 0.008<br>(0.005, 0.011) |
| AIC | 30263.79 | 30263.40 | 30263.79 |
| BIC | 30561.43 | 30577.13 | 30561.43 |
| AUC | 0.660 | 0.661 | 0.660 |
| NB All predictors standardised and centred<br>**AIC** Akaike Information Criterion; **ASES** Arthritis Self-Efficacy Scale; **BIC** Bayesian Information Criterion; **EQ5D** EuroQoL-5 dimensions; **HOOS** Hip Osteoarthritis Outcome Score; **ICC** Intra-class Correlation Coefficient; **IQR** Interquartile range; **KOOS** Knee Osteoarthritis Outcome Score; **MCS** Mental Component Score; **aOR** adjusted Odds Ratio; **PCS** Physical Component Score; **QOL** Quality of Life; **SD** Standard deviation; **UCLA** University of California, Los Angeles; **VAS** Visual Analogue Score; **VPC** Variance Partition Coefficient; **95%CI** 95 percent confidence interval |  |  |  |

Baseline patient characteristics associated (p<0.05) with increased odds of achieving a ≥30% reduction in pain at 3 months were: younger age, female sex, primary complaint was knee problem, lower BMI, shorter duration of complaint, faster walking speed, greater number of chair stands, fewer areas of body pain, higher arthritis self-efficacy scores, higher HOOS/KOOS Quality of Life subscale scores, higher EQ5D VAS scores, lower SF-12 Physical Component Scale scores, higher SF-12 Mental Component Scale scores, and higher pain intensity. Although the AIC remained constant across the three models, the BIC for model 2 was lower than that for model 1, indicating that the addition of therapist-level characteristics did improve model fit. However, therapist-level predictors - cumulative number of patients treated (aOR 0.97 (95%CI: 0.93, 1.02)) and number of days since completing GLA:D training (0.99 (0.94, 1.04)) – were not statistically significantly associated with the outcome. The ICC (0.007) in the null model was statistically significant and effectively unchanged after adjusting for patient-level characteristics (0.008) and therapist-level characteristics (0.008).The AUC of 0.66 suggests weak discriminative ability.

The funnel plot showing therapist-specific outcomes adjusted for patient- and therapist-level covariates and corrected for overdispersion, identified two outlier therapists with better than expected outcomes at the 99.8% confidence threshold (**Figure 1**).

**Figure 1:**
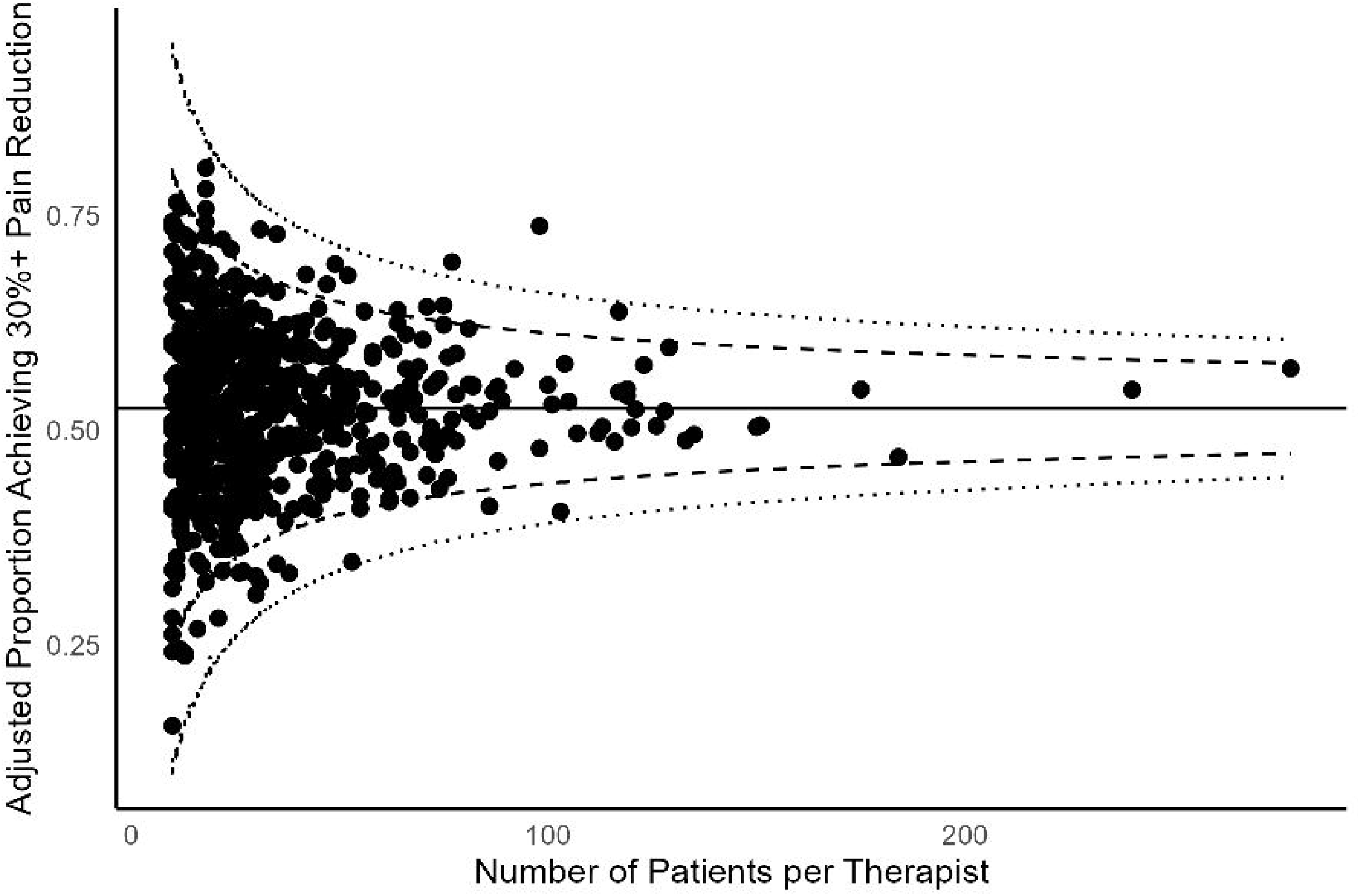
Funnel plot of case-mix adjusted pain relief outcomes by therapist: GLA:D 2014-2019

Across all secondary outcomes and models in the multiply imputed data analyses, the pattern of predictor-outcome associations and the magnitude of the ICC was similar (**Table 5; Supplementary Data**).

**Table 5.** Summary of ‘therapist effects’ for primary and secondary outcomes, multiply imputed data and complete case analysis.

|  | ‘Null’ model | Patient-level adjustment model | ‘Full model’ |
| --- | --- | --- | --- |
|  | N=23,021 | N=23,021 | N=23,021 |
|  | ICC (95%CI) | ICC (95%CI) | ICC (95%CI) |
| **Outcome: Proportion of patients achieving 30% reduction in pain intensity at 3 months** |  |  |  |
| Multiply imputed datasets (n=23,021) | 0.007<br>(0.005, 0.009) | 0.008<br>(0.005, 0.011) | 0.008<br>(0.005, 0.011) |
| Complete case analysis (n=9,720) | 0.014<br>(0.005, 0.026) | 0.015<br>(0.005, 0.025) | 0.015<br>(0.005, 0.024) |
| **Outcome: Proportion of patients achieving 50% reduction in pain intensity at 3 months** |  |  |  |
| Multiply imputed data (n=23,021) | 0.007<br>(0.004, 0.009) | 0.007<br>(0.004, 0.010) | 0.007<br>(0.004, 0.010) |
| Complete case analysis (n=9,720) | 0.012<br>(0.002, 0.022) | 0.013<br>(0.003, 0.024) | 0.013<br>(0.001, 0.022) |
| **Outcome: Pain intensity score (0-100) at 3 months** |  |  |  |
| Multiply imputed data (n=23,021) | 0.008<br>(0.006, 0.010) | 0.009<br>(0.006, 0.011) | 0.009<br>(0.006, 0.011) |
| Complete case analysis (n=9,720) | 0.015<br>(0.006, 0.023) | 0.016<br>(0.005, 0.025) | 0.016<br>(0.007, 0.022) |
| **Outcome: HOOS/KOOS Quality of Life Subscale Score (0-100) at 3 months** |  |  |  |
| Multiply imputed data (n=23,021) | 0.008<br>(0.007, 0.010) | 0.006<br>(0.005, 0.008) | 0.006<br>(0.005, 0.008) |
| Complete case analysis (n=9,720) | 0.014<br>(0.006, 0.026) | 0.012<br>(0.002, 0.022) | 0.012<br>(0.003, 0.022) |
| **Outcome: EQ5D Health Utility Score (-0.757 – 1.000) at 3 months** |  |  |  |
| Multiply imputed data (n=23,021) | 0.008<br>(0.006, 0.010) | 0.003<br>(0.001, 0.005) | 0.003<br>(0.001, 0.005) |
| Complete case analysis (n=9,720) | 0.012<br>(0.001, 0.022) | 0.008<br>(0.001, 0.016) | 0.008<br>(0.000, 0.019) |
| **Outcome: EQ5D VAS Score (0-100) at 3 months** |  |  |  |
| Multiply imputed data (n=23,021) | 0.011 | 0.004 | 0.004 |
|  | (0.009, 0.013) | (0.002, 0.006) | (0.002, 0.005) |
| Complete case analysis (n=9,720) | 0.015<br>(0.001, 0.026) | 0.008<br>(0.001, 0.015) | 0.008<br>(0.002, 0.018) |
| **EQ5D** EuroQoL 5 Dimensions Health-Related Quality of Life; **HOOS/KOOS** Hip Injury and Osteoarthritis Outcome Score/Knee Injury and Osteoarthritis Score; **ICC** Intraclass Correlation Coefficient; **VAS** Visual Analogue Scale |  |  |  |

Complete case analysis included 9720 participants and 403 therapists with complete data on all outcomes and covariates. Across all outcomes and models, the ICC estimates were systematically higher than in the analyses of multiply imputed data. However, the upper 95% confidence limits for all ICC estimates was below 0.03 (**Table 5; Supplementary Data**).

## DISCUSSION

### Summary of key findings

The present study used data available from a large-scale national registry to estimate the magnitude of ‘therapist effects’ on short-term patient-reported pain and quality of life outcomes for adults enrolled on a structured, group-based, physiotherapist-led osteoarthritis management program. Our multi-level models suggested small or very small therapist effects, accounting for less than 3% of the variance across all outcomes, analyses, and models.

### Comparison with previous studies

Our estimates are at the lowest end of the range of 1-12% reported in previous studies of therapist effects in musculoskeletal pain conditions,[10–14] and the 3-12% range typically observed in other conditions and settings[4–7,34]. Before speculating on substantive reasons why this might be, it is important to consider whether this could reflect model mis-specification and unreasonable assumptions during modelling.[35]

Excluding outlier therapists would be expected to reduce therapist effects. Selection of therapists for inclusion in our analyses was based only on having seen a minimum of 10 patients – a suggested minimum for such analyses.[3] We were careful not to select or exclude any therapists based on their characteristics or outcomes, including the number of their patients providing incomplete data and missing outcome data at 3 months. Overall, 27% of eligible participants did not have the primary outcome observed, which was expected to vary by therapist, and could not be assumed to be missing completely at random. In these circumstances complete-case analysis not only loses statistical power but is likely to be susceptible to bias.[36] Our primary analysis was therefore based on multi-level multiply imputed data, including imputing missing outcome data. Our findings assume data missing at random and correct specification of the multiple imputation procedures. We did not find substantially larger estimates from complete case analysis, suggesting that any mis-specification of the imputation procedures may be unlikely to mask substantially higher ICC values. Nevertheless, it is possible that the (self-)selection and training of therapists to deliver a standardised intervention effectively reduces the amount of therapist variance that is present in the data. Indeed, this would be an intentional goal to ensure the GLA:D program is delivered in a consistent manner. In studies of psychotherapy, manualised treatment has been associated with lower therapist effects.[37] Previous studies in musculoskeletal rehabilitation using routine or registry data were largely based on unselected physiotherapists providing an unspecified mixture of treatment approaches under ‘usual care’.[11–14] Lewis et al.[10] provide direct comparisons of ICC values between trial treatment arms, including usual care, but the highly variable ICC estimates likely also reflected limited sample sizes for their stratified analyses.

Inappropriate model assumptions and analytic processes may also inflate patient variation, and consequently reduce the contribution of therapist effects to total variation in outcomes. The choice of outcome measure and timing of endpoint seem unlikely to explain the difference: previous studies have used a mixture of pain/symptom and functional outcomes and endpoints with no obvious relationship to ICC estimates emerging. ICC estimates from two trials using self-reported functional outcome at 3 months were 2.6%[38] and 2.7%[39] respectively. Our study chose a combination of binary and continuous outcomes across three domains, pain, hip/knee-specific quality of life, and health-related quality of life. The magnitude of ICC estimates did not differ substantially from one outcome to another.

### Strengths and Limitations

This the largest study to date of therapist effects in musculoskeletal rehabilitation, made possible by the routine collection and recording of consistent data on patient-reported outcomes and therapists at scale in the nationwide GLA:D program. It comfortably exceeds the suggested guidance of a minimum of 100 therapists each treating at least 10 patients. We were able to incorporate a large number of patient-level covariates in the case-mix adjustment. Higher pain severity, longer symptom duration, multiple site pain, older age, and higher BMI were consistently associated with worse outcomes in the current study. These are well-recognised prognostic indicators of poor outcome in musculoskeletal pain conditions.[40]

There were some limitations in the data available. “Ability to participate in daily activities” is a core outcome domain for osteoarthritis management programs[21] but KOOS-12 Function was introduced only partway through the period covered in the current analysis. Few therapist-level factors were available within the GLA:D registry data, and this meant it was not possible to extend previous work exploring the importance of personality traits and attitudes for therapist effects.[11–14] Outcome measures were available at 12 months, however we reasoned that therapist effects would most likely be observed in the short-term. An alternative hypothesis might be that some therapists are more effective at engaging patients and creating sustained outcomes. Future studies of longer-term outcomes could investigate this. Similarly, it could be of interest to extend our analyses to other outcomes such as functional performance tests. Our choice of 30% reduction in pain intensity for primary analysis is open to challenge. OMERACT-OARSI criteria[41], for example, consider 20% improvement (coupled with improvement in function or patient global assessment) as clinically important improvement. However, our secondary analyses included a higher threshold (50%) and pain intensity as a continuous outcome, and found consistent ICC estimates. Based on this, we would not expect a change to 20% threshold to alter our findings on therapist effects.

GLA:D is delivered as a group intervention, whereas the previous estimates of therapist effects in musculoskeletal rehabilitation have come from individual treatment. If some of the variation in patient outcomes is due to interactions between group members independent of the therapist then this may contribute to the lower estimates of therapist effects in the current study.

### Implications for research and/or practice

Implications of our findings depend to some extent on whether therapist effects are viewed as contextual or specific effects, and whether we believe that selection and training of therapists for GLA:D, together with standardisation of the GLA:D intervention, have effectively minimised the scope for variation in therapist effects on patient outcomes. Therapist effects may be viewed as part of wider concepts of contextual effects, often additive to ‘specific’ treatment effects.[42] Substantial contextual factors are seen across most treatments for osteoarthritis[43,44], and seem likely also for exercise-based interventions.[45] Our findings extend this work by suggesting that contextual factors that relate to therapist effects – therapist characteristics or therapist-patient interaction and alliance[46] - make a minimal contribution to variation in patient outcomes from this structured, group-based rehabilitation intervention. Any contextual effects must be attributable to alternative sources, e.g. patient expectations, intervention setting.

Therapist effects may also be viewed as integral to ‘specific’ treatment effects.[47,48] The recently developed core capability framework for qualified healthcare professionals asserts that health professionals require a diverse array of skills to provide optimal care, including rehabilitative interventions, for all people with OA.[49]

The lack of therapist effects seen in our study could be consistent with two alternative explanations: either these capabilities are fairly consistently present among all therapists delivering the GLA:D intervention, or that patient outcomes from this structured program are not particularly dependent on them. Determining between these competing explanations has important implications for the selection and training of therapists to deliver structured rehabilitation programs, perhaps even the expansion of this role to other personnel. The emergence of osteoarthritis management programs internationally, with varying structure, context, and content, may present opportunities not just for replication of our findings but crucial insights that discriminate between these competing alternative explanations[50]. At the present time, our study suggests that further efforts to select and match therapists to deliver osteoarthritis management programs, or to monitor therapist-level outcomes over and above current practice in GLA:D appear unwarranted.

## Conclusion

National GLA:D registry data provide a unique opportunity to investigate the magnitude of therapist effects on patient outcomes from a structured, group-based osteoarthritis management program. We found that therapist effects – whether viewed as contextual factors or integral to specific treatment effects - made a minimal contribution to variation in patient outcomes.

## Supporting information

STROBE checklist

## Data Availability

The data used in this study cannot be shared publicly because of potentially identifiable or sensitive information. Data may be accessed upon reasonable request by contacting the GLA:D administration.

## Funding

The initiation of the Good Life With Osteoarthritis in Denmark Program was partly funded by the Danish Physiotherapy Association’s fund for research, education, and practice development; the Danish Rheumatism Association; and the Physiotherapy Practice Foundation. STSs research is currently funded by a program grant from Region Zealand (Exercise First), a consortium grant from the European Union’s Horizon 2020 research and innovation program under grant agreement No 945377 (ESCAPE) and an Innovative Health Initiative Joint Undertaking (IHI JU) under grant agreement No. 101219324 (PROBE). DTG is currently funded by a grant from Danish Regions (No R232-A5132, a faculty grant from The Department of Sports Science and Clinical Biomechanics, Faculty of Health Sciences, University of Southern Denmark, and a grant from NSR Hospitals, Denmark (No A1683), which all are outside the submitted work. Data access costs were funded by an award from the School of Health and Social Care, Sheffield Hallam University. GMP is part funded by the EPSRC South Yorkshire Digital Health Hub (EP/X03075X/1).

## Ethical approval

The GLA:D® registry was approved by the Danish Data Protection Agency. According to the Danish Data Protection Act patient consent is not required as personal data was processed exclusively for research and statistical purposes. Separate ethics approval was not needed for the current analysis.

## Contributorship

**PEO:** formal analysis, methodology, software, visualization, writing – original draft, writing – review and editing; **JP:** conceptualization, methodology, writing – original draft, writing – review and editing; **DA:** conceptualization, methodology, writing – original draft, writing – review and editing; **DY:** conceptualisation, methodology, writing – review and editing; **DTG:** Data curation, investigation, methodology, project management, resources, writing – review and editing; **EMR:** Funding acquisition, investigation, methodology, supervision, writing – review and editing; **STS:** Funding acquisition, investigation, methodology, supervision, writing – review and editing; **GMP:** conceptualisation, formal analysis, funding acquisition, methodology, project management, software, supervision, validation, visualization, writing – original draft, writing – review and editing

## Competing interests

EMR is the copyright holder of Knee injury and Osteoarthritis Outcome Score (KOOS) and several other patient-reported outcome measures, and co-founder of the Good Life with Osteoarthritis in Denmark (GLA:D®), a not-for profit initiative to implement clinical guidelines in primary care hosted by University of Southern Denmark. STS has received personal fees from Munksgaard and TrustMe-Ed, outside the submitted work, and is co-founder of GLA:D®. PEO, DA, JP, DY, DTG, GMP have no competing interests to declare.

## Data Sharing

The data used in this study cannot be shared publicly because of potentially identifiable or sensitive information. Data may be accessed upon reasonable request by contacting the GLA:D® administration.

## Acknowledgements

The authors gratefully acknowledge the contribution of Professor Khaled Khatab to the design, analysis, and interpretation of the work. For the purpose of open access, the author has applied a Creative Commons Attribution (CC BY) licence to any Author Accepted Manuscript version arising from this submission.

## Notes

### Funding Statement

This study was funded by the Danish Physiotherapy Association, the Danish Rheumatism Association, the Physiotherapy Practice Foundation, Region Zealand (Exercise First), EU Horizon 2020 (No 945377 (ESCAPE)), Innovative Health Initiative Joint Undertaking (IHI JU) (No. 101219324 (PROBE)), Danish Regions (No R232-A5132), The Department of Sports Science and Clinical Biomechanics, Faculty of Health Sciences, University of Southern Denmark, NSR Hospitals, Denmark (No A1683), School of Health and Social Care, Sheffield Hallam University, and the EPSRC South Yorkshire Digital Health Hub (EP/X03075X/1).

### Author Declarations

The Danish Data Protection Agency waived ethical approval for this work.

## REFERENCES

1 Thomas J, Petticrew M, Noyes J, et al. Chapter 17: Intervention complexity. In: Higgins J, Thomas J, Chandler J, et al., eds. Cochrane Handbook for Systematic Reviews of Interventions. Cochrane 2023.

2 Lambert M. Bergin and Garfield’s handbook of psychotherapy and behaviour change. 6th ed. Hoboken: John Wiley & Sons 2013.

3 Johns RG, Barkham M, Kellett S, et al. A systematic review of therapist effects: A critical narrative update and refinement to review. Clin Psychol Rev. 2019;67:78–93. doi: 10.1016/j.cpr.2018.08.004

4 Kim D-M, Wampold BE, Bolt DM. Therapist effects in psychotherapy: A random-effects modeling of the National Institute of Mental Health Treatment of Depression Collaborative Research Program data. Psychotherapy Research. 2006;16:161–72. doi: 10.1080/10503300500264911

5 Schnelle C, Clark J, Mascord R, et al. Is There a Surgeons’ Effect on Patients’ Physical Health, Beyond the Intervention, That Requires Further Investigation? A Systematic Review. Ther. Clin. Risk Manag. 2022;18:467–90. doi: 10.2147/TCRM.S357934

6 Schnelle C, Clark J, Mascord R, et al. Is There a Doctors’ Effect on Patients’ Physical Health, Beyond the Intervention and All Known Factors? A Systematic Review. Ther. Clin. Risk Manag. 2022;18:721–37. doi: 10.2147/TCRM.S372464

7 Berglar J, Crameri A, Von Wyl A, et al. Therapist effects on treatment outcome in psychotherapy: a multilevel modelling analysis. International Journal of Psychotherapy. 2016;20:61–80.

8 Soldati S, Colais P, Davoli M, et al. More is more? The role of surgeon in the volume-outcome relationship: an Italian population-based cohort study. BMJ Open . 2025;15. doi: 10.1136/bmjopen-2024-098569

9 Anderson T, McClintock AS, Himawan L, et al. A Prospective Study of Therapist Facilitative Interpersonal Skills as a Predictor of Treatment Outcome. J Consult Clin Psychol. 2016;84:57–66. doi: 10.1037/ccp0000060

10 Lewis M, Morley S, Van Der Windt DAWM, et al. Measuring practitioner/therapist effects in randomised trials of low back pain and neck pain interventions in primary care settings. European Journal of Pain. 2010;14:1033–9. doi: 10.1016/j.ejpain.2010.04.002

11 Kooijman MK, Buining EM, Swinkels ICS, et al. Do therapist effects determine outcome in patients with shoulder pain in a primary care physiotherapy setting? Physiotherapy (United Kingdom). 2020;107:111–7. doi: 10.1016/j.physio.2019.08.009

12 Buining EM, Kooijman MK, Swinkels ICS, et al. Exploring physiotherapists’ personality traits that may influence treatment outcome in patients with chronic diseases: a cohort study. BMC Health Serv Res. 2015;15:558. doi: 10.1186/s12913-015-1225-1

13 Simon CB, Stryker SE, George SZ. Assessing the influence of treating therapist and patient prognostic factors on recovery from axial pain. Journal of Manual and Manipulative Therapy. 2013;21:187–95. doi: 10.1179/2042618613Y.0000000035

14 Nudelman Y, Pincus T, Ami N Ben. Association Between Physical Therapists’ Attitudes and Beliefs and the Functional Outcomes of Patients With Low Back Pain: A Multilevel Analysis Study. Phys Ther. 2025;105. doi: 10.1093/ptj/pzaf007

15 Skou ST, Roos EM. Good Life with osteoArthritis in Denmark (GLA:D™): evidence-based education and supervised neuromuscular exercise delivered by certified physiotherapists nationwide. BMC Musculoskelet Disord. 2017;18:1–13. doi: 10.1186/s12891-017-1439-y

16 McAlindon TE, Driban JB, Henrotin Y, et al. OARSI Clinical Trials Recommendations: Design, conduct, and reporting of clinical trials for knee osteoarthritis. Osteoarthritis Cartilage. 2015;23:747–60. doi: 10.1016/j.joca.2015.03.005

17 Smith SM, Dworkin RH, Turk DC, et al. Interpretation of chronic pain clinical trial outcomes: IMMPACT recommended considerations. Pain. 2020;161:2446–61. doi: 10.1097/j.pain.0000000000001952

18 Dworkin RH, Turk DC, Wyrwich KW, et al. Interpreting the Clinical Importance of Treatment Outcomes in Chronic Pain Clinical Trials: IMMPACT Recommendations. J Pain. 2008;9:105–21. doi: 10.1016/j.jpain.2007.09.005

19 Bellamy N, Kirwan J, Boers M, et al. Recommendations for a Core Set of Outcome Measures for Future Phase III Clinical Trials in Knee, Hip, and Hand Osteoarthritis. Consensus Development at OMERACT III. J Rheumatol 1997;24(4):799–802. PMID: 9101522

20 Smith TO, Hawker GA, Hunter DJ, et al. The OMERACT-OARSI Core Domain Set for Measurement in Clinical Trials of Hip and/or Knee Osteoarthritis. J Rheumatol. 2019;46:981–9. doi: 10.3899/jrheum.181194

21 Allen KD, Huffman K, Cleveland RJ, et al. Evaluating Osteoarthritis Management Programs: outcome domain recommendations from the OARSI Joint Effort Initiative. Osteoarthritis Cartilage. 2023;31:954–65. doi: 10.1016/j.joca.2023.02.078

22 Burgess R, Lewis M, Hill JC. Benchmarking community/primary care musculoskeletal services: A narrative review and recommendation. Musculoskeletal Care. 2023;21:148–58. doi: 10.1002/msc.1676

23 Baumbach L, List M, Grønne DT, et al. Individualized predictions of changes in knee pain, quality of life and walking speed following patient education and exercise therapy in patients with knee osteoarthritis – a prognostic model study. Osteoarthritis Cartilage. 2020;28:1191–201. doi: 10.1016/j.joca.2020.05.014

24 Johnsen MB, Roos E, Grønne DT, et al. Impact of educational level and employment status on short-term and long-term pain relief from supervised exercise therapy and education: an observational study of 22D588 patients with knee and hip osteoarthritis. BMJ Open. 2021;11:e045156. doi: 10.1136/bmjopen-2020-045156

25 Pihl K, Roos EM, Taylor RS, et al. Associations between comorbidities and immediate and one-year outcomes following supervised exercise therapy and patient education – A cohort study of 24,513 individuals with knee or hip osteoarthritis. Osteoarthritis Cartilage. 2021;29:39–49. doi: 10.1016/j.joca.2020.11.001

26 Peat G, Yu D, Grønne DT, et al. Do Patients With Intersectional Disadvantage Have Poorer Outcomes From Osteoarthritis Management Programs? A Tapered Balancing Study of Patient Outcomes From the Good Life With Osteoarthritis in Denmark Program. Arthritis Care Res (Hoboken). 2023;75:136–44. doi: 10.1002/acr.24987

27 Sterne JAC, White IR, Carlin JB, et al. Multiple imputation for missing data in epidemiological and clinical research: potential and pitfalls. BMJ. 2009;338:b2393–b2393. doi: 10.1136/bmj.b2393

28 Rubin DB. Multiple Imputation for Nonresponse in Surveys. John Wiley & Sons, Inc., New York, 1987. doi:10.1002/9780470316696

29 Little RJA, Rubin DB. Statistical Analysis with Missing Data. John Wiley & Sons, Inc., New York, 2002. doi:10.1002/9781119013563

30 Kontopantelis E, White IR, Sperrin M, et al. Outcome-sensitive multiple imputation: A simulation study. BMC Med Res Methodol. 2017;17. doi: 10.1186/s12874-016-0281-5

31 White IR, Royston P, Wood AM. Multiple imputation using chained equations: Issues and guidance for practice. Stat Med. 2011;30:377–99. doi: 10.1002/sim.4067

32 Spiegelhalter DJ. Funnel plots for comparing institutional performance. Stat Med. 2005;24:1185–202. doi: 10.1002/sim.1970

33 Spiegelhalter DJ. Handling over-dispersion of performance indicators. Qual Saf Health Care. 2005;14:347–51. doi: 10.1136/qshc.2005.013755

34 Johns RG, Barkham M, Kellett S, et al. A systematic review of therapist effects: A critical narrative update and refinement to Baldwin and Imel’s (2013) review. Clin. Psychol. Rev. 2019;67:78–93. doi: 10.1016/j.cpr.2018.08.004

35 Wampold BE, Bolt DM. Therapist effects: Clever ways to make them (and everything else) disappear. Psychotherapy Research. 2006;16:184–7. doi: 10.1080/10503300500265181

36 Hughes RA, Heron J, Sterne JAC, et al. Accounting for missing data in statistical analyses: Multiple imputation is not always the answer. Int J Epidemiol. 2019;48:1294–304. doi: 10.1093/ije/dyz032

37 Crits-Christoph P, Baranackie K, Kurcias J, et al. MetaDAnalysis of Therapist Effects in Psychotherapy Outcome Studies. Psychotherapy Research. 1991;1:81–91. doi: 10.1080/10503309112331335511

38 Jellema P, van der Windt DAWM, van der Horst HE, et al. Should treatment of (sub)acute low back pain be aimed at psychosocial prognostic factors? Cluster randomised clinical trial in general practice. BMJ. 2005;331:84. doi: 10.1136/bmj.38495.686736.E0

39 Hay E, Mullis R, Lewis M, et al. Comparison of physical treatments versus a brief pain-management programme for back pain in primary care: a randomised clinical trial in physiotherapy practice. The Lancet. 2005;365:2024–30. doi: 10.1016/S0140-6736(05)66696-2

40 Mallen CD, Peat G, Thomas E, et al. Prognostic factors for musculoskeletal pain in primary care: a systematic review. Br J Gen Pract. 2007;57:655–61. doi: 10.1136/bmjopen-2016-012901

41 Pham T, van der Heijde D, Altman RD, et al. OMERACT-OARSI Initiative: Osteoarthritis Research Society International set of responder criteria for osteoarthritis clinical trials revisited. Osteoarthritis Cartilage. 2004;12:389–99. doi: 10.1016/j.joca.2004.02.001

42 Saueressig T, Pedder H, Owen PJ, et al. Contextual effects: how to, and how not to, quantify them. BMC Med. Res. Methodol. 2024;24. doi: 10.1186/s12874-024-02152-2

43 Zhang W, Robertson J, Jones AC, et al. The placebo effect and its determinants in osteoarthritis: Meta-analysis of randomised controlled trials. Ann Rheum Dis. 2008;67:1716–23. doi: 10.1136/ard.2008.092015

44 Zou K, Wong J, Abdullah N, et al. Examination of overall treatment effect and the proportion attributable to contextual effect in osteoarthritis: Meta-Analysis of randomised controlled trials. Ann Rheum Dis. 2016;75:1964–70. doi: 10.1136/annrheumdis-2015-208387

45 de Roode A, Heymans MW, van Lankveld W, et al. The impact of contextual effects in exercise therapy for low back pain: a systematic review and meta-analysis. BMC Med. 2024;22. doi: 10.1186/s12916-024-03679-3

46 Cook CE, Bailliard A, Bent JA, et al. An international consensus definition for contextual factors: findings from a nominal group technique. Front Psychol. 2023;14. doi: 10.3389/fpsyg.2023.1178560

47 Zilcha-Mano S, Roose SP, Brown PJ, et al. Not just nonspecific factors: The roles of alliance and expectancy in treatment, and their neurobiological underpinnings. Front. Behav. Neurosci. 2019;12:293. doi: 10.3389/fnbeh.2018.00293

48 Stulberg JJ, Huang R, Kreutzer L, et al. Association Between Surgeon Technical Skills and Patient Outcomes. JAMA Surg. 2020;155:960. doi: 10.1001/jamasurg.2020.3007

49 Hinman RS, Allen KD, Bennell KL, et al. Development of a core capability framework for qualified health professionals to optimise care for people with osteoarthritis: an OARSI initiative. Osteoarthritis Cartilage. 2020;28:154–66. doi: 10.1016/j.joca.2019.12.001

50 Platt JR. Strong Inference: Certain systematic methods of scientific thinking may produce much more rapid progress than others. Science 1964;146(3642):347–53. doi: 10.1126/science.146.3642.347

