## Supplementary data for "Therapist effects in real-world rehabilitation outcomes: a cohort study of the nationwide GLA:D osteoarthritis management program in Denmark"

| List of R packages used | 2 |
| --- | --- |
| **Figure S1.** Heatmap of missing data | 4 |
| **Table S1.** Descriptive characteristics of patients (n=23,021) and therapists (n=657) after multiple imputation | 5 |
| **Table S2.** Descriptive characteristics of patients (n=9,720) and therapists (n=403) in the complete case analysis dataset | 6 |
| **Model coefficients and performance on multiply imputed data for secondary outcomes** |  |
| **Table S3.** ≥50% pain reduction at 3 months | 7 |
| **Table S4.** Pain intensity (0-100) at 3 months | 9 |
| **Table S5.** HOOS/KOOS Quality of Life Score (0-100) at 3 months | 10 |
| **Table S6.** EQ5D Health Utility Score (-0.757-1.000) at 3 months | 12 |
| **Table S7.** EQ5D VAS (0-100) at 3 months | 13 |
| **Model coefficients and performance on complete case analysis for primary and secondary outcomes** |  |
| **Table S8.** ≥50% pain reduction at 3 months | 14 |
| **Table S9.** Pain intensity (0-100) at 3 months | 16 |
| **Table S10.** HOOS/KOOS Quality of Life Score (0-100) at 3 months | 17 |
| **Table S11.** EQ5D Health Utility Score (-0.757-1.000) at 3 months | 19 |
| **Table S12.** EQ5D VAS (0-100) at 3 months | 20 |
| **Table S13.** ≥30% pain reduction at 3 months | 21 |
| **Funnel plots of therapist-specific case-mix-adjusted outcomes on multiply imputed data** |  |
| **Figure S2.** ≥50% pain reduction at 3 months | 24 |
| **Figure S3.** Pain intensity (0-100) at 3 months | 24 |
| **Figure S4.** HOOS/KOOS Quality of Life Score (0-100) at 3 months | 25 |
| **Figure S5.** EQ5D Health Utility Score (-0.757-1.000) at 3 months | 25 |
| **Figure S6.** EQ5D VAS (0-100) at 3 months | 26 |
| **Funnel plots of therapist-specific case-mix-adjusted outcomes on complete case analysis** |  |
| **Figure S7.** ≥50% pain reduction at 3 months | 26 |
| **Figure S8.** Pain intensity (0-100) at 3 months | 27 |
| **Figure S9.** HOOS/KOOS Quality of Life Score (0-100) at 3 months | 27 |
| **Figure S10.** EQ5D Health Utility Score (-0.757-1.000) at 3 months | 28 |
| **Figure S11.** EQ5D VAS (0-100) at 3 months | 28 |
| **Figure S12.** ≥30% pain reduction at 3 months | 29 |
| **Figure S13.** Summary of ICC estimates of ‘therapist effects’ for primary and secondary outcomes, multiply imputed data and complete case analysis | 30 |

**List of R packages used**

| boot | Canty A, Ripley B (2025). _boot: Bootstrap Functions_. R package version 1.3-32, <https://CRAN.R-project.org/package=boot>.  Davison A, Hinkley D (1997). _Bootstrap Methods and Their Applications_. Cambridge University Press, Cambridge. ISBN 0-521-57391-2, doi:10.1017/CBO9780511802843 <https://doi.org/10.1017/CBO9780511802843>. |
| --- | --- |
| broom | Robinson D, Hayes A, Couch S (2025). _broom: Convert Statistical Objects into Tidy Tibbles_. R package version 1.0.10, <https://CRAN.R-project.org/package=broom>. |
| car | Fox J, Weisberg S (2019). _An R Companion to Applied Regression_, Third edition. Sage, Thousand Oaks CA.<https://www.john-fox.ca/Companion/>. |
| DescTools | Signorell A (2025). _DescTools: Tools for Descriptive Statistics_. R package version 0.99.60, <https://CRAN.R-project.org/package=DescTools>. |
| DHARMa | Hartig F (2024). _DHARMa: Residual Diagnostics for Hierarchical (Multi-Level / Mixed) Regression Models_. R package version 0.4.7, <https://CRAN.R-project.org/package=DHARMa>. |
| dplyr | Wickham H, François R, Henry L, Müller K, Vaughan D (2023). _dplyr: A Grammar of Data Manipulation_. R package version 1.1.4, <https://CRAN.R-project.org/package=dplyr>. |
| ggplot2 | H. Wickham. ggplot2: Elegant Graphics for Data Analysis. Springer-Verlag New York, 2016. |
| haven | Wickham H, Miller E, Smith D (2025). _haven: Import and Export 'SPSS', 'Stata' and 'SAS' Files_. R package version 2.5.5, <https://CRAN.R-project.org/package=haven>. |
| Hmisc | Harrell Jr F (2025). _Hmisc: Harrell Miscellaneous_. R package version 5.2-4, <https://CRAN.R-project.org/package=Hmisc>. |
| lme4 | Douglas Bates, Martin Maechler, Ben Bolker, Steve Walker (2015). Fitting Linear Mixed-Effects Models Using lme4. Journal of Statistical Software, 67(1), 1-48. doi:10.18637/jss.v067.i01. |
| lmerTest | Kuznetsova A, Brockhoff PB, Christensen RHB (2017). “lmerTest Package: Tests in Linear Mixed Effects Models.” _Journal of Statistical Software_, *82*(13), 1-26. doi:10.18637/jss.v082.i13 <https://doi.org/10.18637/jss.v082.i13>. |
| mice | Stef van Buuren, Karin Groothuis-Oudshoorn (2011). mice: Multivariate Imputation by Chained Equations in R. Journal of Statistical |
| miceadds | Robitzsch, A., & Grund, S. (2025). miceadds: Some Additional Multiple Imputation Functions, Especially for 'mice'. R package version 3.18-36. <https://CRAN.R-project.org/package=miceadds> Software, 45(3), 1-67. DOI 10.18637/jss.v045.i03. |
| mitml | Grund S, Robitzsch A, Luedtke O (2023). _mitml: Tools for Multiple Imputation in Multilevel Modeling_. R package version 0.4-5, <https://CRAN.R-project.org/package=mitml>. |
| narian | Tierney N, Cook D (2023). “Expanding Tidy Data Principles to Facilitate Missing Data Exploration, Visualization and Assessment of Imputations.” _Journal of Statistical Software_, *105*(7), 1-31. doi:10.18637/jss.v105.i07 <https://doi.org/10.18637/jss.v105.i07>. |
| openxlsx | Schauberger P, Walker A (2025). _openxlsx: Read, Write and Edit xlsx Files_. R package version 4.2.8.1, <https://CRAN.R-project.org/package=openxlsx>. |
| performance | Lüdecke et al., (2021). performance: An R Package for Assessment, Comparison and Testing of Statistical Models. Journal of Open Source Software, 6(60), 3139. https://doi.org/10.21105/joss.03139 |
| pROC | Xavier Robin, Natacha Turck, Alexandre Hainard, Natalia Tiberti, Frédérique Lisacek, Jean-Charles Sanchez and Markus Müller (2011). pROC: an open-source package for R and S+ to analyze and compare ROC curves. BMC Bioinformatics, 12, p. 77. DOI: 10.1186/1471-2105-12-77 <http://www.biomedcentral.com/1471-2105/12/77/> |
| readxl | Wickham H, Bryan J (2025). _readxl: Read Excel Files_. R package version 1.4.5, <https://CRAN.R-project.org/package=readxl>. |
| scales | Wickham H, Pedersen T, Seidel D (2025). _scales: Scale Functions for Visualization_. R package version 1.4.0. <https://CRAN.R-project.org/package=scales>. |
| sjPlot | Lüdecke D (2025). _sjPlot: Data Visualization for Statistics in Social Science_. R package version 2.9.0, <https://CRAN.R-project.org/package=sjPlot>. |
| skimr | Waring E, Quinn M, McNamara A, Arino de la Rubia E, Zhu H, Ellis S (2025). _skimr: Compact and Flexible Summaries of Data_. R package version 2.2.1, <https://CRAN.R-project.org/package=skimr>. |
| stringr | Wickham H (2025). _stringr: Simple, Consistent Wrappers for Common String Operations_. R package version 1.6.0, <https://CRAN.R-project.org/package=stringr>. |
| tidyr | Wickham H, Vaughan D, Girlich M (2024). _tidyr: Tidy Messy Data_. R package version 1.3.1, <https://CRAN.R-project.org/package=tidyr>. |
| VIM | Alexander Kowarik, Matthias Templ (2016). Imputation with the R Package VIM. Journal of Statistical Software, 74(7), 1-16. doi:10.18637/jss.v074.i07 |
| writexl | Ooms J (2025). _writexl: Export Data Frames to Excel 'xlsx' Format_. R package version 1.5.4, <https://CRAN.R-project.org/package=writexl>. |

**Figure S1.** Heatmap of missing data
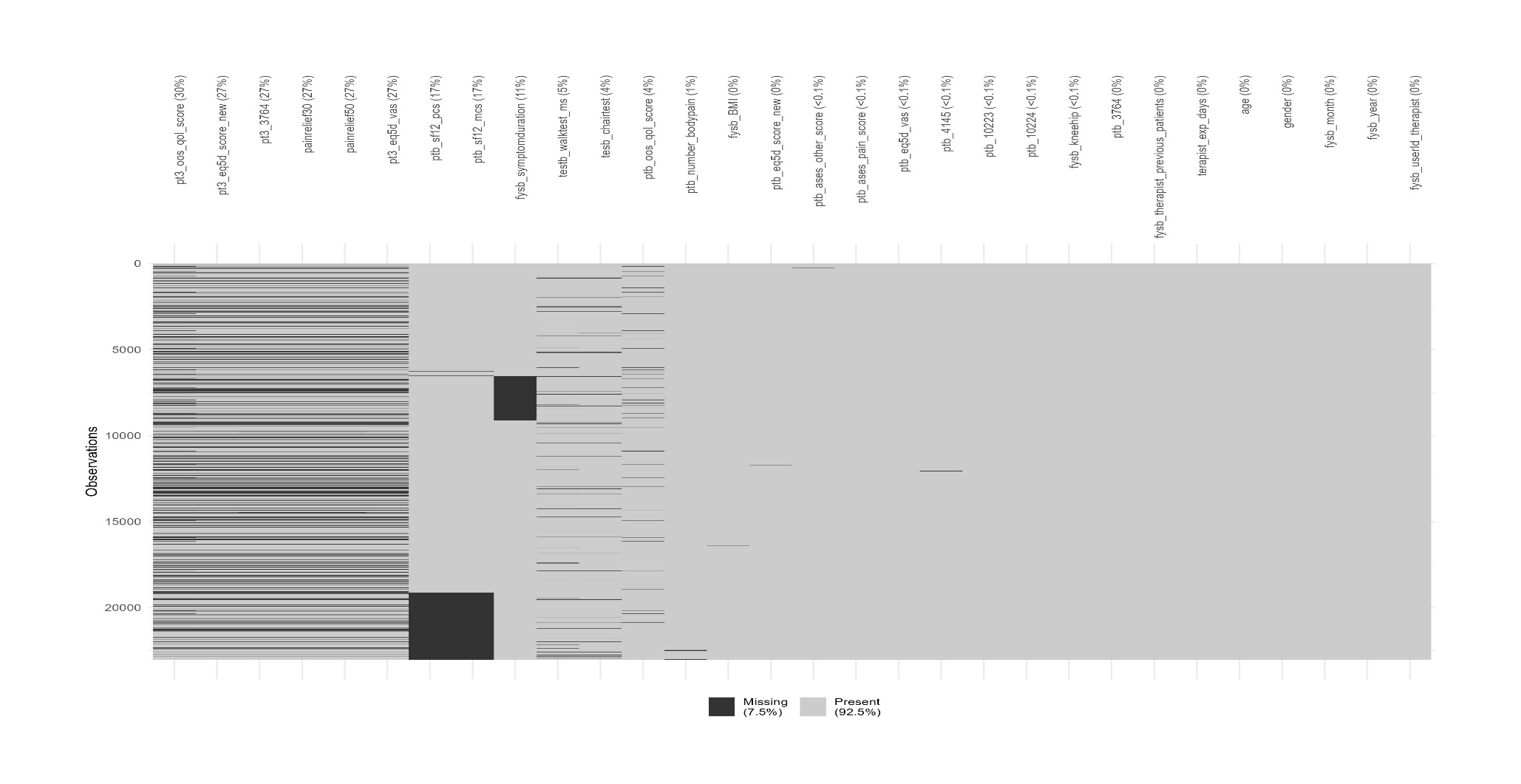

**Table S1.** Descriptive characteristics of patients (n=23,021) and therapists (n=657) after multiple imputation

|  | **Valid N** | **Miss %** | **%** |
| --- | --- | --- | --- |
| **Therapists** | | | |
| Number of patients treated: median (IQR) | 657 | *0* | 36 (16, 72) |
| Days since therapist certification: median (IQR) | 657 | *0* | 645 (313, 1077) |
| **Patients** | | | |
| Age (years): mean (SD) | 23,021 | *0* | 65.0 (9.8) |
| Female sex | 23,021 | *0* | *74.4* |
| Born in Denmark | 23,009 | *<1* | *96.0* |
| Danish citizen | 23,009 | *<1* | *98.4* |
| Body mass index (kg/m^2^): mean (SD) | 22,956 | *<1* | 29.0 (5.5) |
| Symptom duration (months): median (IQR) | 20,408 | *11* | 18 (6, 48) |
| 40m walk test at baseline (m/sec): mean (SD) | 21,792 | *5* | 1.4 (0.3) |
| Chair test at baseline (no.): mean (SD) | 22,158 | *4* | 11.4 (3.8) |
| No. of pain areas at baseline: mean (SD) | 22,851 | *<1* | 4.0 (3.3) |
| UCLA Activity score (0-10): mean (SD) | 23,004 | *<1* | 5.5 (1.8) |
| Primary complaint | 23,019 | *<1* |  |
| Hip |  |  | *25.8* |
| Knee |  |  | *74.2* |
| Month of enrollment on GLA:D | 23,021 | *0* |  |
| January |  |  | *11.4* |
| February |  |  | *7.6* |
| March |  |  | *8.3* |
| April |  |  | *8.6* |
| May |  |  | *7.6* |
| June |  |  | *5.7* |
| July |  |  | *4.4* |
| August |  |  | *12.6* |
| September |  |  | *9.6* |
| October |  |  | *11.8* |
| November |  |  | *7.0* |
| December |  |  | *5.4* |
| Year of enrollment on GLA:D | 23,021 | *0* |  |
| 2014 |  |  | *5.3* |
| 2015 |  |  | *14.5* |
| 2016 |  |  | *22.4* |
| 2017 |  |  | *21.6* |
| 2018 |  |  | *19.5* |
| 2019 |  |  | *16.8* |
| Arthritis Self-Efficacy Scale: Pain at baseline (10-100): mean (SD) | 23,002 | *<1* | 61.9 (20.0) |
| Arthritis Self-Efficacy Scale: Other at baseline (10-100): mean (SD) | 22,998 | *<1* | 66.6 (18.3) |
| SF-12 Physical Component Summary at baseline (0-100): mean (SD) | 19,076 | *17* | 35.3 (8.2) |
| SF-12 Mental Component Summary at baseline (0-100): mean (SD) | 19,076 | *17* | 51.1 (10.1) |
| Pain intensity at baseline (0-100): mean (SD) | 23,021 | *0* | 61.3 (14.5) |
| Pain intensity at 3 months (0-100): mean (SD) | 16,746 | *27* | 41.2 (22.6) |
| Experienced ≥30% reduction in pain at 3 months | 16,746 | *27* | *52.6* |
| Experienced ≥50% reduction in pain at 3 months | 16,746 | *27* | *36.9* |
| HOOS/KOOS QOL at baseline (0-100): mean (SD) | 22,215 | *4* | 40.9 (14.3) |
| HOOS/KOOS QOL at 3 months (0-100): mean (SD) | 16,221 | *30* | 47.5 (16.5) |
| EQ5D VAS at baseline (0-100): mean (SD) | 23,003 | *<1* | 64.8 (19.0) |
| EQ5D VAS at 3 months (0-100): mean (SD) | 16,748 | *27* | 69.2 (19.3) |
| EQ5D utility score at baseline (-0.757-1.000): mean (SD) | 22,996 | *<1* | 0.710 (0.216) |
| EQ5D utility score at 3 months (-0.757-1.000): mean (SD) | 16,743 | *27* | 0.774 (0.209) |
| Figures represent N (%) unless otherwise stated  Miss % = rate of missing data for each variable  **GLA:D** Good Life with osteoArthritis in Denmark; **EQ5D** EuroQoL-5 dimensions; **HOOS** Hip Osteoarthritis Outcome Score; **IQR** Interquartile range; **KOOS** Knee Osteoarthritis Outcome Score; **QOL** Quality of Life; **SD** Standard deviation; **UCLA** University of California, Los Angeles; **VAS** Visual Analogue Score | | | |

**Table S2.** Descriptive characteristics of patients (n=9,720) and therapists (n=403) in the complete case analysis dataset

|  | **Valid N** | **Miss %** | **N** | **%** |
| --- | --- | --- | --- | --- |
| **Therapists** |  |  |  |  |
| Number of patients treated: median (IQR) | 403 | *0* | 18 (13, 29) | |
| Days since therapist certification: median (IQR) | 403 | *0* | 591 (304, 995) | |
| **Patients** |  |  |  |  |
| Age (years): mean (SD) | 9,720 | *0* | 64.8 (9.5) | |
| Female sex | 9,720 | *0* | 7,297 | *75.1* |
| Born in Denmark | 9,720 | *0* | 9,370 | *96.4* |
| Danish citizen | 9,720 | *0* | 9,575 | *98.5* |
| Body mass index (kg/m^2^): mean (SD) | 9,720 | *0* | 28.8 (5.4) | |
| Symptom duration (months): median (IQR) | 9,720 | *0* | 18 (6, 50) | |
| 40m walk test at baseline (m/sec): mean (SD) | 9,720 | *0* | 1.5 (0.3) | |
| Chair test at baseline (no.): mean (SD) | 9,720 | *0* | 11.6 (3.7) | |
| No. of pain areas at baseline: mean (SD) | 9,720 | *0* | 4.6 (3.0) | |
| UCLA Activity score (0-10): mean (SD) | 9,720 | *0* | 5.6 (1.8) | |
| Primary complaint | 9,720 | *0* |  | |
| Hip |  |  | 2,418 | *24.9* |
| Knee |  |  | 7,302 | *75.1* |
| Month of enrollment on GLA:D | 9,720 | *0* |  |  |
| January |  |  | 1,255 | *12.9* |
| February |  |  | 809 | *8.3* |
| March |  |  | 879 | *9.0* |
| April |  |  | 958 | *9.9* |
| May |  |  | 681 | *7.0* |
| June |  |  | 420 | *4.3* |
| July |  |  | 363 | *3.7* |
| August |  |  | 1063 | *10.9* |
| September |  |  | 743 | *7.6* |
| October |  |  | 963 | *9.9* |
| November |  |  | 822 | *8.5* |
| December |  |  | 764 | *7.9* |
| Year of enrollment on GLA:D | 9,720 | *0* |  | |
| 2014 |  |  | 792 | *8.1* |
| 2015 |  |  | 2,192 | *22.6* |
| 2016 |  |  | 1,565 | *16.1* |
| 2017 |  |  | 2,854 | *29.4* |
| 2018 |  |  | 2,317 | *23.8* |
| 2019 |  |  | 0 | *0.0* |
| Arthritis Self-Efficacy Scale: Pain at baseline (10-100): mean (SD) | 9,720 | *0* | 63.5 (19.6) | |
| Arthritis Self-Efficacy Scale: Other at baseline (10-100): mean (SD) | 9,720 | *0* | 68.1 (17.6) | |
| SF-12 Physical Component Summary at baseline (0-100): mean (SD) | 9,720 | *0* | 35.7 (8.2) | |
| SF-12 Mental Component Summary at baseline (0-100): mean (SD) | 9,720 | *0* | 51.6 (9.9) | |
| Pain intensity at baseline (0-100): mean (SD) | 9,720 | *0* | 60.8 (14.2) | |
| Pain intensity at 3 months (0-100): mean (SD) | 9,720 | *0* | 41.0 (22.6) | |
| Experienced ≥30% reduction in pain at 3 months | 9,720 | *0* | 5,150 | *53.0* |
| Experienced ≥50% reduction in pain at 3 months | 9,720 | *0* | 3,442 | *35.4* |
| HOOS/KOOS QOL at baseline (0-100): mean (SD) | 9,720 | *0* | 41.2 (13.9) | |
| HOOS/KOOS QOL at 3 months (0-100): mean (SD) | 9,720 | *0* | 47.6 (16.5) | |
| EQ5D VAS at baseline (0-100): mean (SD) | 9,720 | *0* | 66.1 (18.4) | |
| EQ5D VAS at 3 months (0-100): mean (SD) | 9,720 | *0* | 70.3 (18.7) | |
| EQ5D utility score at baseline (-0.757-1.000): mean (SD) | 9,720 | *0* | 0.723 (0.205) | |
| EQ5D utility score at 3 months (-0.757-1.000): mean (SD) | 9,720 | *0* | 0.781 (0.201) | |
| Figures represent N (%) unless otherwise stated  Miss % = rate of missing data for each variable  **GLA:D** Good Life with osteoArthritis in Denmark; **EQ5D** EuroQoL-5 dimensions; **HOOS** Hip Osteoarthritis Outcome Score; **IQR** Interquartile range; **KOOS** Knee Osteoarthritis Outcome Score; **QOL** Quality of Life; **SD** Standard deviation; **UCLA** University of California, Los Angeles; **VAS** Visual Analogue Score | | | | |

**Table S3.** Random intercept logistic regression models for ≥50% pain reduction at 3 months: multiply imputed data

|  | Model 0 | Model 1 | Model 2 |
| --- | --- | --- | --- |
|  | ‘Null’ model | Patient-level adjustment model | ‘Full model’ |
|  | N=23,021 | N=23,021 | N=23,021 |
|  |  | aOR (95%CI) | aOR (95%CI) |
| **Therapist-level variables** |  |  |  |
| Number of patients treated |  |  | 0.96 (0.92, 1.01) |
| Days since therapist certification |  |  | 1.01 (0.96, 1.07) |
| **Patient-level variables** |  |  |  |
| Age |  | **0.93 (0.90, 0.97)** | **0.93 (0.90, 0.97)** |
| Female sex |  | **1.08 (1.00, 1.17)** | **1.08 (1.00, 1.17)** |
| Born in Denmark |  | 1.03 (0.84, 1.28) | 1.03 (0.84, 1.28) |
| Danish citizen |  | 1.20 (0.87, 1.65) | 1.20 (0.87, 1.65) |
| Month of entry (ref: January) |  |  |  |
| February |  | 1.07 (0.92, 1.25) | 1.07 (0.92, 1.25) |
| March |  | **1.28 (1.11, 1.48)** | **1.29 (1.11, 1.49)** |
| April |  | **1.15 (1.00, 1.32)** | **1.15 (1.00, 1.32)** |
| May |  | **1.31 (1.12, 1.52)** | **1.31 (1.13, 1.53)** |
| June |  | **1.21 (1.04, 1.42)** | **1.22 (1.04, 1.43)** |
| July |  | 1.04 (0.87, 1.25) | 1.05 (0.88, 1.25) |
| August |  | 1.13 (0.98, 1.29) | 1.13 (0.98, 1.30) |
| September |  | 1.13 (0.98, 1.30) | 1.13 (0.98, 1.30) |
| October |  | **1.19 (1.05, 1.36)** | **1.19 (1.05, 1.36)** |
| November |  | 1.15 (0.99, 1.35) | **1.16 (1.00, 1.35)** |
| December |  | 1.08 (0.91, 1.29) | 1.09 (0.92, 1.30) |
| Year of entry (ref: 2014) |  |  |  |
| 2015 |  | 0.92 (0.79, 1.08) | 0.93 (0.80, 1.09) |
| 2016 |  | **0.85 (0.73, 0.99)** | 0.86 (0.74, 1.01) |
| 2017 |  | 1.02 (0.88, 1.19) | 1.04 (0.89, 1.22) |
| 2018 |  | 1.08 (0.92, 1.26) | 1.10 (0.93, 1.30) |
| 2019 |  | 0.93 (0.79, 1.09) | 0.96 (0.80, 1.15) |
| Most affected joint is hip (ref: knee) |  | **0.84 (0.78, 0.90)** | **0.84 (0.78, 0.90)** |
| BMI (kg/m^2^) |  | **0.95 (0.92, 0.99)** | **0.95 (0.92, 0.99)** |
| Duration of symptoms (months) |  | **0.89 (0.86, 0.93)** | **0.89 (0.86, 0.93)** |
| 40m walk test at baseline |  | **1.09 (1.04, 1.14)** | **1.09 (1.04, 1.14)** |
| No. of chair stands during 30sec at baseline |  | **1.05 (1.01, 1.09)** | **1.05 (1.01, 1.09)** |
| No. of painful body areas (0-56) at baseline |  | **0.89 (0.86, 0.93)** | **0.89 (0.86, 0.93)** |
| UCLA - Physical activity (1-10) at baseline |  | 0.98 (0.95, 1.02) | 0.98 (0.95, 1.02) |
| KOOS/HOOS QOL (0-100) at baseline |  | **1.10 (1.05, 1.14)** | **1.09 (1.05, 1.14)** |
| ASES: Other (10-100) at baseline |  | 0.98 (0.92, 1.04) | 0.98 (0.92, 1.04) |
| ASES: Pain (10-100) at baseline |  | **1.26 (1.19, 1.33)** | **1.26 (1.19, 1.33)** |
| EQ-VAS (0-100) at baseline |  | **1.09 (1.04, 1.14)** | **1.09 (1.04, 1.14)** |
| EQ-5D Health Utility score (-0.757-1) at baseline |  | **1.19 (1.13, 1.25)** | **1.19 (1.13, 1.25)** |
| SF-12 PCS (0-100) at baseline |  | 0.96 (0.92, 1.01) | 0.96 (0.92, 1.01) |
| SF-12 MCS (0-100) at baseline |  | **0.93 (0.89, 0.98)** | **0.93 (0.89, 0.98)** |
| Pain intensity (0-100) at baseline |  | **1.53 (1.48, 1.59)** | **1.53 (1.48, 1.59)** |
| VPC (ICC) | 0.007  (0.004, 0.009) | 0.007  (0.004, 0.010) | 0.007  (0.004, 0.010) |
| AIC | 30301.68 | 28969.16 | 28969.17 |
| BIC | 30317.77 | 29266.79 | 29282.9 |
| AUC | 0.597 | 0.655 | 0.655 |
| NB All predictors standardised and centred  **AIC** Akaike Information Criterion; **ASES** Arthritis Self-Efficacy Scale; **BIC** Bayesian Information Criterion; **EQ5D** EuroQoL-5 dimensions; **HOOS** Hip Osteoarthritis Outcome Score; **ICC** Intra-class Correlation Coefficient; **IQR** Interquartile range; **KOOS** Knee Osteoarthritis Outcome Score; **MCS** Mental Component Score; **aOR** adjusted Odds Ratio; **PCS** Physical Component Score; **QOL** Quality of Life; **SD** Standard deviation; **UCLA** University of California, Los Angeles; **VAS** Visual Analogue Score; **VPC** Variance Partition Coefficient; **95%CI** 95 percent confidence interval | | | |

**Table S4.** Random intercept linear regression models for pain intensity (0-100) at 3 months: multiply imputed data

|  | Model 0 | Model 1 | Model 2 |
| --- | --- | --- | --- |
|  | ‘Null’ model | Patient-level adjustment model | ‘Full model’ |
|  | N=23,021 | N=23,021 | N=23,021 |
|  |  | Adj.b (95%CI) | Adj.b (95%CI) |
| **Therapist-level variables** |  |  |  |
| Number of patients treated |  |  | 0.019 (-0.002, 0.039) |
| Days since therapist certification |  |  | -0.001 (-0.022, 0.020) |
| **Patient-level variables** |  |  |  |
| Age |  | **0.034 (0.020, 0.048)** | **0.034 (0.019, 0.048)** |
| Female sex |  | **-0.037 (-0.065, -0.008)** | **-0.037 (-0.066, -0.009)** |
| Born in Denmark |  | -0.017 (-0.092, 0.058) | -0.016 (-0.091, 0.059) |
| Danish citizen |  | -0.031 (-0.147, 0.085) | -0.031 (-0.147, 0.085) |
| Month of entry (ref: January) |  |  |  |
| February |  | 0.006 (-0.051, 0.062) | 0.006 (-0.051, 0.063) |
| March |  | **-0.056 (-0.112, -0.001)** | **-0.057 (-0.113, -0.002)** |
| April |  | -0.046 (-0.101, 0.009) | -0.047 (-0.101, 0.008) |
| May |  | **-0.070 (-0.127, -0.013)** | **-0.071 (-0.128, -0.014)** |
| June |  | -0.052 (-0.114, 0.011) | -0.055 (-0.117, 0.008) |
| July |  | -0.013 (-0.082, 0.055) | -0.016 (-0.085, 0.052) |
| August |  | -0.033 (-0.083, 0.017) | -0.035 (-0.086, 0.015) |
| September |  | -0.016 (-0.070, 0.037) | -0.020 (-0.073, 0.034) |
| October |  | **-0.070 (-0.121, -0.019)** | **-0.070 (-0.125, -0.023)** |
| November |  | **-0.065 (-0.124, -0.006)** | **-0.069 (-0.128, -0.010)** |
| December |  | 0.024 (-0.040, 0.088) | 0.019 (-0.045, 0.083) |
| Year of entry (ref: 2014) |  |  |  |
| 2015 |  | 0.050 (-0.012, 0.113) | 0.044 (-0.019, 0.107) |
| 2016 |  | **0.094 (0.034, 0.155)** | **0.083 (0.021, 0.145)** |
| 2017 |  | 0.023 (-0.038, 0.084) | 0.007 (-0.058, 0.072) |
| 2018 |  | -0.001 (-0.063, 0.061) | -0.022 (-0.091, 0.047) |
| 2019 |  | 0.043 (-0.021, 0.107) | 0.017 (-0.057, 0.092) |
| Most affected joint is hip (ref: knee) |  | **0.121 (0.093, 0.150)** | **0.121 (0.093, 0.149)** |
| BMI (kg/m^2^) |  | **0.026 (0.013, 0.039)** | **0.026 (0.013, 0.039)** |
| Duration of symptoms (months) |  | **0.051 (0.039, 0.064)** | **0.051 (0.039, 0.063)** |
| 40m walk test at baseline |  | **-0.042 (-0.059, -0.025)** | **-0.043 (-0.060, -0.026)** |
| No. of chair stands during 30sec at baseline |  | **-0.026 (-0.040, -0.011)** | **-0.026 (-0.041, -0.011)** |
| No. of painful body areas (0-56) at baseline |  | **0.044 (0.031, 0.057)** | **0.044 (0.031, 0.057)** |
| UCLA - Physical activity (1-10) at baseline |  | 0.009 (-0.004, 0.023) | 0.009 (-0.004, 0.023) |
| KOOS/HOOS QOL (0-100) at baseline |  | **-0.050 (-0.066, -0.035)** | **-0.050 (-0.066, -0.035)** |
| ASES: Other (10-100) at baseline |  | 0.012 (-0.009, 0.033) | 0.012 (-0.009, 0.033) |
| ASES: Pain (10-100) at baseline |  | **-0.111 (-0.135, -0.097)** | **-0.116 (-0.136, -0.097)** |
| EQ-VAS (0-100) at baseline |  | **-0.049 (-0.064, -0.033)** | **-0.048 (-0.064, -0.033)** |
| EQ-5D Health Utility score (-0.757-1) at baseline |  | **-0.111 (-0.129, -0.094)** | **-0.111 (-0.128, -0.094)** |
| SF-12 PCS (0-100) at baseline |  | 0.003 (-0.014, 0.020) | 0.003 (-0.014, 0.020) |
| SF-12 MCS (0-100) at baseline |  | 0.031 (0.016, 0.046) | 0.031 (0.016, 0.046) |
| Pain intensity (0-100) at baseline |  | **0.119 (0.105, 0.133)** | **0.119 (0.105, 0.133)** |
| VPC (ICC) | 0.008  (0.006, 0.010) | 0.009  (0.006, 0.011) | 0.009  (0.006, 0.011) |
| AIC | 65172.45 | 62549.8 | 62563.48 |
| BIC | 65196.58 | 62855.48 | 62885.24 |
| NB All predictors standardised and centred  **Adj.b** Adjusted beta coefficient; **AIC** Akaike Information Criterion; **ASES** Arthritis Self-Efficacy Scale; **BIC** Bayesian Information Criterion; **EQ5D** EuroQoL-5 dimensions; **HOOS** Hip Osteoarthritis Outcome Score; **ICC** Intra-class Correlation Coefficient; **IQR** Interquartile range; **KOOS** Knee Osteoarthritis Outcome Score; **MCS** Mental Component Score; **PCS** Physical Component Score; **QOL** Quality of Life; **SD** Standard deviation; **UCLA** University of California, Los Angeles; **VAS** Visual Analogue Score; **VPC** Variance Partition Coefficient; **95%CI** 95 percent confidence interval | | | |

**Table S5.** Random intercept linear regression models for HOOS/KOOS Quality of Life Score (0-100) at 3 months: multiply imputed data

|  | Model 0 | Model 1 | Model 2 |
| --- | --- | --- | --- |
|  | ‘Null’ model | Patient-level adjustment model | ‘Full model’ |
|  | N=23,021 | N=23,021 | N=23,021 |
|  |  | Adj.b (95%CI) | Adj.b (95%CI) |
| **Therapist-level variables** |  |  |  |
| Number of patients treated |  |  | -0.005 (-0.022, 0.012) |
| Days since therapist certification |  |  | 0.016 (-0.003, 0.034) |
| **Patient-level variables** |  |  |  |
| Age |  | **0.029 (0.017, 0.042)** | **0.030 (0.017, 0.042)** |
| Female sex |  | **0.060 (0.034, 0.085)** | **0.060 (0.034, 0.085)** |
| Born in Denmark |  | 0.004 (-0.066, 0.067) | 0.001 (-0.066, 0.068) |
| Danish citizen |  | -0.027 (-0.130, 0.077) | -0.026 (-0.129, 0.077) |
| Month of entry (ref: January) |  |  |  |
| February |  | -0.006 (-0.057, 0.044) | -0.005 (-0.056, 0.045) |
| March |  | 0.046 (-0.004, 0.095) | 0.045 (-0.004, 0.094) |
| April |  | **0.053 (0.004, 0.102)** | **0.052 (0.003, 0.101)** |
| May |  | 0.038 (-0.013, 0.089) | 0.037 (-0.013, 0.088) |
| June |  | **0.060 (0.005, 0.116)** | **0.059 (0.003, 0.115)** |
| July |  | 0.015 (-0.046, 0.076) | 0.014 (-0.047, 0.075) |
| August |  | 0.040 (-0.004, 0.085) | 0.036 (-0.009, 0.081) |
| September |  | 0.036 (-0.012, 0.084) | 0.033 (-0.015, 0.080) |
| October |  | **0.052 (0.007, 0.097)** | **0.048 (0.003, 0.093)** |
| November |  | 0.036 (-0.017, 0.089) | 0.032 (-0.020, 0.085) |
| December |  | -0.035 (-0.092, 0.023) | -0.039 (-0.096, 0.018) |
| Year of entry (ref: 2014) |  |  |  |
| 2015 |  | -0.001 (-0.056, 0.055) | -0.006 (-0.062, 0.050) |
| 2016 |  | -0.011 (-0.064, 0.043) | -0.021 (-0.076, 0.034) |
| 2017 |  | -0.024 (-0.078, 0.031) | -0.039 (-0.096, 0.019) |
| 2018 |  | 0.027 (-0.028, 0.082) | 0.005 (-0.055, 0.066) |
| 2019 |  | 0.017 (-0.039, 0.074) | -0.010 (-0.076, 0.055) |
| Most affected joint is hip (ref: knee) |  | -0.012 (-0.037, 0.014) | -0.012 (-0.037, 0.013) |
| BMI (kg/m^2^) |  | **-0.013 (-0.025, -0.001)** | **-0.013 (-0.025, -0.001)** |
| Duration of symptoms (months) |  | **-0.045 (-0.056, -0.034)** | **-0.045 (-0.056, -0.034)** |
| 40m walk test at baseline |  | **0.019 (0.004, 0.033)** | **0.019 (0.004, 0.034)** |
| No. of chair stands during 30sec at baseline |  | **0.027 (0.014, 0.040)** | **0.026 (0.013, 0.039)** |
| No. of painful body areas (0-56) at baseline |  | -0.022 (-0.033, -0.010) | -0.022 (-0.033, -0.010) |
| UCLA - Physical activity (1-10) at baseline |  | **-0.025 (-0.037, -0.013)** | **-0.025 (-0.037, -0.013)** |
| KOOS/HOOS QOL (0-100) at baseline |  | **0.364 (0.350, 0.378)** | **0.364 (0.350, 0.378)** |
| ASES: Other (10-100) at baseline |  | 0.005 (-0.014, 0.024) | 0.005 (-0.014, 0.024) |
| ASES: Pain (10-100) at baseline |  | **0.082 (0.065, 0.099)** | **0.082 (0.065, 0.100)** |
| EQ-VAS (0-100) at baseline |  | **0.028 (0.014, 0.042)** | **0.028 (0.014, 0.042)** |
| EQ-5D Health Utility score (-0.757-1) at baseline |  | **0.072 (0.056, 0.087)** | **0.072 (0.056, 0.087)** |
| SF-12 PCS (0-100) at baseline |  | **0.100 (0.085, 0.115)** | **0.100 (0.085, 0.115)** |
| SF-12 MCS (0-100) at baseline |  | **0.026 (0.013, 0.040)** | **0.026 (0.013, 0.040)** |
| Pain intensity (0-100) at baseline |  | **0.014 (0.001, 0.026)** | **0.014 (0.001, 0.026)** |
| VPC (ICC) | 0.008  (0.007, 0.010) | 0.006  (0.005, 0.008) | 0.006  (0.005, 0.008) |
| AIC | 65185.74 | 57259.65 | 57275.56 |
| BIC | 65209.87 | 57565.32 | 57597.33 |
| NB All predictors standardised and centred  **Adj.b** Adjusted beta coefficient; **AIC** Akaike Information Criterion; **ASES** Arthritis Self-Efficacy Scale; **BIC** Bayesian Information Criterion; **EQ5D** EuroQoL-5 dimensions; **HOOS** Hip Osteoarthritis Outcome Score; **ICC** Intra-class Correlation Coefficient; **IQR** Interquartile range; **KOOS** Knee Osteoarthritis Outcome Score; **MCS** Mental Component Score; **PCS** Physical Component Score; **QOL** Quality of Life; **SD** Standard deviation; **UCLA** University of California, Los Angeles; **VAS** Visual Analogue Score; **VPC** Variance Partition Coefficient; **95%CI** 95 percent confidence interval | | | |

**Table S6.** Random intercept linear regression models for EQ5D Health Utility Score (-0.757-1.000) at 3 months: multiply imputed data

|  | Model 0 | Model 1 | Model 2 |
| --- | --- | --- | --- |
|  | ‘Null’ model | Patient-level adjustment model | ‘Full model’ |
|  | N=23,021 | N=23,021 | N=23,021 |
|  |  | Adj.b (95%CI) | Adj.b (95%CI) |
| **Therapist-level variables** |  |  |  |
| Number of patients treated |  |  | -0.008 (-0.024, 0.008) |
| Days since therapist certification |  |  | 0.003 (-0.015, 0.020) |
| **Patient-level variables** |  |  |  |
| Age |  | **0.016 (0.003, 0.029)** | **0.016 (0.004, 0.029)** |
| Female sex |  | **0.124 (0.099, 0.150)** | **0.125 (0.099, 0.150)** |
| Born in Denmark |  | **-0.076 (-0.143, -0.008)** | **-0.076 (-0.143, -0.008)** |
| Danish citizen |  | -0.024 (-0.128, 0.080) | -0.024 (-0.128, 0.080) |
| Month of entry (ref: January) |  |  |  |
| February |  | 0.001 (-0.050, 0.052) | 0.001 (-0.050, 0.052) |
| March |  | 0.004 (-0.046, 0.053) | 0.004 (-0.045, 0.054) |
| April |  | 0.013 (-0.036, 0.062) | 0.013 (-0.036, 0.062) |
| May |  | 0.013 (-0.038, 0.065) | 0.014 (-0.037, 0.065) |
| June |  | 0.031 (-0.025, 0.087) | 0.032 (-0.024, 0.088) |
| July |  | 0.050 (-0.011, 0.111) | 0.051 (-0.010, 0.112) |
| August |  | 0.027 (-0.017, 0.072) | 0.027 (-0.017, 0.072) |
| September |  | 0.015 (-0.033, 0.063) | 0.016 (-0.032, 0.064) |
| October |  | **0.059 (0.014, 0.104)** | **0.060 (0.014, 0.105)** |
| November |  | 0.028 (-0.025, 0.081) | 0.029 (-0.024, 0.082) |
| December |  | -0.011 (-0.068, 0.047) | -0.009 (-0.067, 0.048) |
| Year of entry (ref: 2014) |  |  |  |
| 2015 |  | -0.023 (-0.079, 0.033) | -0.022 (-0.078, 0.035) |
| 2016 |  | -0.019 (-0.072, 0.035) | -0.017 (-0.071, 0.038) |
| 2017 |  | -0.025 (-0.079, 0.030) | -0.022 (-0.078, 0.035) |
| 2018 |  | -0.039 (-0.094, 0.016) | -0.035 (-0.095, 0.024) |
| 2019 |  | -0.046 (-0.102, 0.011) | -0.041 (-0.105, 0.023) |
| Most affected joint is hip (ref: knee) |  | **-0.141 (-0.167, -0.116)** | **-0.141 (-0.167, -0.116)** |
| BMI (kg/m^2^) |  | **-0.013 (-0.025, -0.001)** | **-0.013 (-0.025, -0.001)** |
| Duration of symptoms (months) |  | **-0.018 (-0.029, -0.007)** | **-0.018 (-0.029, -0.007)** |
| 40m walk test at baseline |  | **0.039 (0.024, 0.053)** | **0.039 (0.024, 0.054)** |
| No. of chair stands during 30sec at baseline |  | **0.018 (0.005, 0.031)** | **0.018 (0.005, 0.031)** |
| No. of painful body areas (0-56) at baseline |  | **-0.030 (-0.041, -0.018)** | **-0.030 (-0.041, -0.018)** |
| UCLA - Physical activity (1-10) at baseline |  | 0.005 (-0.007, 0.017) | 0.005 (-0.007, 0.017) |
| KOOS/HOOS QOL (0-100) at baseline |  | **0.059 (0.045, 0.073)** | **0.059 (0.045, 0.073)** |
| ASES: Other (10-100) at baseline |  | **0.047 (0.028, 0.065)** | **0.047 (0.028, 0.066)** |
| ASES: Pain (10-100) at baseline |  | **0.053 (0.035, 0.070)** | **0.053 (0.036, 0.071)** |
| EQ-VAS (0-100) at baseline |  | **0.044 (0.030, 0.057)** | **0.044 (0.030, 0.057)** |
| EQ-5D Health Utility score (-0.757-1) at baseline |  | **0.355 (0.339, 0.370)** | **0.355 (0.339, 0.370)** |
| SF-12 PCS (0-100) at baseline |  | **0.063 (0.048, 0.078)** | **0.063 (0.048, 0.078)** |
| SF-12 MCS (0-100) at baseline |  | **0.068 (0.054, 0.082)** | **0.068 (0.055, 0.082)** |
| Pain intensity (0-100) at baseline |  | -0.004 (-0.017, 0.008) | -0.004 (-0.017, 0.008) |
| VPC (ICC) | 0.008  (0.006, 0.010) | 0.003  (0.001, 0.005) | 0.003  (0.001, 0.005) |
| AIC | 66094.29 | 57553.07 | 57571.25 |
| BIC | 66118.43 | 57858.75 | 57893.02 |
| NB All predictors standardised and centred  **Adj.b** Adjusted beta coefficient; **AIC** Akaike Information Criterion; **ASES** Arthritis Self-Efficacy Scale; **BIC** Bayesian Information Criterion; **EQ5D** EuroQoL-5 dimensions; **HOOS** Hip Osteoarthritis Outcome Score; **ICC** Intra-class Correlation Coefficient; **IQR** Interquartile range; **KOOS** Knee Osteoarthritis Outcome Score; **MCS** Mental Component Score; **PCS** Physical Component Score; **QOL** Quality of Life; **SD** Standard deviation; **UCLA** University of California, Los Angeles; **VAS** Visual Analogue Score; **VPC** Variance Partition Coefficient; **95%CI** 95 percent confidence interval | | | |

**Table S7.** Random intercept linear regression models for EQ5D VAS (0-100) at 3 months: multiply imputed data

|  | Model 0 | Model 1 | Model 2 |
| --- | --- | --- | --- |
|  | ‘Null’ model | Patient-level adjustment model | ‘Full model’ |
|  | N=23,021 | N=23,021 | N=23,021 |
|  |  | Adj.b (95%CI) | Adj.b (95%CI) |
| **Therapist-level variables** |  |  |  |
| Number of patients treated |  |  | -0.001 (-0.017, 0.016) |
| Days since therapist certification |  |  | **0.018 (0.001, 0.036)** |
| **Patient-level variables** |  |  |  |
| Age |  | 0.010 (-0.003, 0.023) | 0.010 (-0.003, 0.023) |
| Female sex |  | **0.052 (0.026, 0.077)** | **0.052 (0.026, 0.077)** |
| Born in Denmark |  | 0.039 (-0.029, 0.106) | 0.039 (-0.028, 0.107) |
| Danish citizen |  | 0.006 (-0.098, 0.110) | 0.007 (-0.097, 0.111) |
| Month of entry (ref: January) |  |  |  |
| February |  | 0.019 (-0.032, 0.070) | 0.020 (-0.031, 0.071) |
| March |  | -0.003 (-0.053, 0.046) | -0.004 (-0.054, 0.046) |
| April |  | 0.028 (-0.021, 0.077) | 0.027 (-0.022, 0.077) |
| May |  | 0.019 (-0.032, 0.071) | 0.019 (-0.033, 0.070) |
| June |  | 0.011 (-0.046, 0.067) | 0.008 (-0.048, 0.065) |
| July |  | -0.033 (-0.094, 0.028) | -0.036 (-0.097, 0.026) |
| August |  | 0.040 (-0.005, 0.085) | 0.034 (-0.011, 0.079) |
| September |  | -0.002 (-0.050, 0.046) | -0.007 (-0.055, 0.041) |
| October |  | -0.012 (-0.058, 0.033) | -0.018 (-0.064, 0.028) |
| November |  | 0.014 (-0.039, 0.067) | 0.009 (-0.044, 0.063) |
| December |  | 0.007 (-0.050, 0.065) | 0.001 (-0.057, 0.059) |
| Year of entry (ref: 2014) |  |  |  |
| 2015 |  | -0.025 (-0.081, 0.031) | -0.033 (-0.089, 0.024) |
| 2016 |  | **-0.055 (-0.108, -0.001)** | **-0.068 (-0.123, -0.013)** |
| 2017 |  | **-0.065 (-0.120, -0.011)** | **-0.086 (-0.143, -0.029)** |
| 2018 |  | **-0.070 (-0.125, -0.015)** | **-0.099 (-0.159, -0.039)** |
| 2019 |  | **-0.125 (-0.182, -0.068)** | **-0.162 (-0.227, -0.098)** |
| Most affected joint is hip (ref: knee) |  | **-0.096 (-0.122, -0.071)** | **-0.097 (-0.122, -0.071)** |
| BMI (kg/m^2^) |  | **-0.024 (-0.036, -0.012)** | **-0.024 (-0.036, -0.012)** |
| Duration of symptoms (months) |  | **-0.024 (-0.035, -0.013)** | **-0.024 (-0.035, -0.013)** |
| 40m walk test at baseline |  | **0.022 (0.008, 0.037)** | **0.023 (0.008, 0.037)** |
| No. of chair stands during 30sec at baseline |  | **0.022 (0.009, 0.035)** | **0.022 (0.008, 0.035)** |
| No. of painful body areas (0-56) at baseline |  | **-0.041 (-0.052, -0.029)** | **-0.041 (-0.052, -0.029)** |
| UCLA - Physical activity (1-10) at baseline |  | **0.021 (0.008, 0.033)** | **0.021 (0.008, 0.032)** |
| KOOS/HOOS QOL (0-100) at baseline |  | **0.018 (0.004, 0.032)** | **0.018 (0.003, 0.032)** |
| ASES: Other (10-100) at baseline |  | **0.087 (0.068, 0.106)** | **0.087 (0.068, 0.106)** |
| ASES: Pain (10-100) at baseline |  | **0.054 (0.036, 0.071)** | **0.053 (0.036, 0.071)** |
| EQ-VAS (0-100) at baseline |  | **0.286 (0.272, 0.300)** | **0.286 (0.272, 0.300)** |
| EQ-5D Health Utility score (-0.757-1) at baseline |  | **0.036 (0.020, 0.51)** | **0.036 (0.020, 0.51)** |
| SF-12 PCS (0-100) at baseline |  | **0.116 (0.101, 0.131)** | **0.116 (0.101, 0.131)** |
| SF-12 MCS (0-100) at baseline |  | **0.083 (0.069, 0.097)** | **0.083 (0.069, 0.096)** |
| Pain intensity (0-100) at baseline |  | 0.009 (-0.003, 0.021) | 0.009 (-0.004, 0.021) |
| VPC (ICC) | 0.011  (0.009, 0.013) | 0.004  (0.002, 0.006) | 0.004  (0.002, 0.005) |
| AIC | 65465.27 | 57694.32 | 57707.77 |
| BIC | 65489.40 | 58000.00 | 58029.53 |
| NB All predictors standardised and centred  **Adj.b** Adjusted beta coefficient; **AIC** Akaike Information Criterion; **ASES** Arthritis Self-Efficacy Scale; **BIC** Bayesian Information Criterion; **EQ5D** EuroQoL-5 dimensions; **HOOS** Hip Osteoarthritis Outcome Score; **ICC** Intra-class Correlation Coefficient; **IQR** Interquartile range; **KOOS** Knee Osteoarthritis Outcome Score; **MCS** Mental Component Score; **PCS** Physical Component Score; **QOL** Quality of Life; **SD** Standard deviation; **UCLA** University of California, Los Angeles; **VAS** Visual Analogue Score; **VPC** Variance Partition Coefficient; **95%CI** 95 percent confidence interval | | | |

**Table S8.** Random intercept logistic regression models for ≥50% pain reduction at 3 months: complete case analysis

|  | Model 0 | Model 1 | Model 2 |
| --- | --- | --- | --- |
|  | ‘Null’ model | Patient-level adjustment model | ‘Full model’ |
|  | N=9,720 | N=9,720 | N=9,720 |
|  |  | aOR (95%CI) | aOR (95%CI) |
| **Therapist-level variables** |  |  |  |
| Number of patients treated |  |  | 0.94 (0.88, 1.01) |
| Days since therapist certification |  |  | 1.02 (0.95, 1.10) |
| **Patient-level variables** |  |  |  |
| Age |  | **0.92 (0.87, 0.97)** | **0.92 (0.88, 0.97)** |
| Female sex |  | **1.19 (1.07, 1.32)** | **1.19 (1.08, 1.33)** |
| Born in Denmark |  | 1.02 (0.76, 1.37) | 1.02 (0.76, 1.37) |
| Danish citizen |  | 1.29 (0.83, 2.01) | 1.29 (0.83, 2.01) |
| Month of enrollment on GLA:D (ref: January) |  |  |  |
| February |  | 1.04 (0.86, 1.27) | 1.04 (0.86, 1.27) |
| March |  | **1.38 (1.14, 1.66)** | **1.38 (1.15, 1.67)** |
| April |  | 1.15 (0.96, 1.39) | 1.16 (0.96, 1.39) |
| May |  | **1.32 (1.08, 1.62)** | **1.32 (1.08, 1.62)** |
| June |  | 1.11 (0.87, 1.42) | 1.11 (0.87, 1.43) |
| July |  | 1.11 (0.86, 1.44) | 1.12 (0.86, 1.45) |
| August |  | 1.11 (0.93, 1.34) | 1.11 (0.93, 1.34) |
| September |  | 1.13 (0.92, 1.39) | 1.14 (0.93, 1.40) |
| October |  | **1.22 (1.01, 1.48)** | **1.23 (1.02, 1.49)** |
| November |  | 1.12 (0.92, 1.37) | 1.14 (0.93, 1.38) |
| December |  | 1.10 (0.90, 1.35) | 1.12 (0.92, 1.37) |
| Year of enrollment on GLA:D (ref: 2014) |  |  |  |
| 2015 |  | 1.03 (0.86, 1.24) | 1.05 (0.87, 1.26) |
| 2016 |  | 0.99 (0.81, 1.22) | 1.02 (0.82, 1.25) |
| 2017 |  | 1.09 (0.91, 1.30) | 1.12 (0.92, 1.37) |
| 2018 |  | 1.18 (0.98, 1.42) | 1.23 (0.99, 1.54) |
| 2019 |  | - | - |
| Most affected joint is hip (ref: knee) |  | 0.90 (0.81, 1.00) | 0.90 (0.81, 1.00) |
| BMI (kg/m^2^) |  | 0.95 (0.91, 1.00) | 0.95 (0.91, 1.00) |
| Duration of symptoms (months) |  | **0.88 (0.84, 0.93)** | **0.88 (0.84, 0.93)** |
| 40m walk test at baseline |  | **1.12 (1.05, 1.19)** | **1.12 (1.06, 1.19)** |
| No. of chair stands during 30sec at baseline |  | **1.08 (1.02, 1.14)** | **1.08 (1.02, 1.14)** |
| No. of painful body areas (0-56) at baseline |  | **0.84 (0.80, 0.89)** | **0.84 (0.80, 0.89)** |
| UCLA - Physical activity (1-10) at baseline |  | 0.99 (0.95, 1.04) | 0.99 (0.95, 1.04) |
| KOOS/HOOS QOL (0-100) at baseline |  | **1.14 (1.08, 1.21)** | **1.14 (1.08, 1.21)** |
| ASES: Other (10-100) at baseline |  | 1.01 (0.93, 1.09) | 1.01 (0.93, 1.09) |
| ASES: Pain (10-100) at baseline |  | **1.31 (1.22, 1.40)** | **1.31 (1.22, 1.41)** |
| EQ-VAS (0-100) at baseline |  | **1.07 (1.01, 1.13)** | **1.07 (1.01, 1.13)** |
| EQ-5D Health Utility score (-0.757-1) at baseline |  | **1.15 (1.08, 1.22)** | **1.15 (1.08, 1.22)** |
| SF-12 PCS (0-100) at baseline |  | 0.94 (0.89, 1.00) | 0.94 (0.89, 1.00) |
| SF-12 MCS (0-100) at baseline |  | 0.96 (0.91, 1.02) | 0.96 (0.91, 1.02) |
| Pain intensity (0-100) at baseline |  | **1.34 (1.28, 1.41)** | **1.34 (1.28, 1.41)** |
| VPC (ICC) | 0.012  (0.002, 0.022) | 0.013  (0.003, 0.024) | 0.013  (0.001, 0.022) |
| AIC | 12627.76 | 12065.79 | 12066.7 |
| BIC | 12642.12 | 12324.34 | 12339.62 |
| AUC | 0.622 | 0.667 | 0.667 |
| NB All predictors standardised and centred  **AIC** Akaike Information Criterion; **ASES** Arthritis Self-Efficacy Scale; **BIC** Bayesian Information Criterion; **EQ5D** EuroQoL-5 dimensions; **HOOS** Hip Osteoarthritis Outcome Score; **ICC** Intra-class Correlation Coefficient; **IQR** Interquartile range; **KOOS** Knee Osteoarthritis Outcome Score; **MCS** Mental Component Score; **aOR** adjusted Odds Ratio; **PCS** Physical Component Score; **QOL** Quality of Life; **SD** Standard deviation; **UCLA** University of California, Los Angeles; **VAS** Visual Analogue Score; **VPC** Variance Partition Coefficient; **95%CI** 95 percent confidence interval | | | |

**Table S9.** Random intercept linear regression models for pain intensity (0-100) at 3 months: complete case analysis

|  | Model 0 | Model 1 | Model 2 |
| --- | --- | --- | --- |
|  | ‘Null’ model | Patient-level adjustment model | ‘Full model’ |
|  | N=9,720 | N=9,720 | N=9,720 |
|  |  | Adj.b (95%CI) | Adj.b (95%CI) |
| **Therapist-level variables** |  |  |  |
| Number of patients treated |  |  | 0.026 (-0.005, 0.058) |
| Days since therapist certification |  |  | -0.007 (-0.040, 0.026) |
| **Patient-level variables** |  |  |  |
| Age |  | **0.035 (0.014, 0.056)** | **0.034 (0.013, 0.056)** |
| Female sex |  | **-0.086 (-0.129, -0.042)** | **-0.087 (-0.130, -0.043)** |
| Born in Denmark |  | -0.040 (-0.163, 0.082) | -0.040 (-0.163, 0.083) |
| Danish citizen |  | -0.056 (-0.245, 0.132) | -0.057 (-0.245, 0.131) |
| Month of entry (ref: January) |  |  |  |
| February |  | 0.044 (-0.037, 0.126) | 0.044 (-0.037, 0.126) |
| March |  | -0.085 (-0.164, -0.005) | -0.086 (-0.165, -0.006) |
| April |  | -0.040 (-0.117, 0.038) | -0.040 (-0.118, 0.038) |
| May |  | -0.069 (-0.156, 0.017) | -0.070 (-0.157, 0.016) |
| June |  | -0.033 (-0.136, 0.071) | -0.036 (-0.139, 0.068) |
| July |  | -0.041 (-0.150, 0.069) | -0.044 (-0.153, 0.066) |
| August |  | -0.008 (-0.085, 0.070) | -0.009 (-0.087, 0.069) |
| September |  | -0.025 (-0.110, 0.061) | -0.029 (-0.115, 0.058) |
| October |  | -0.065 (-0.145, 0.015) | -0.068 (-0.149, 0.012) |
| November |  | **-0.087 (-0.169, -0.004)** | **-0.092 (-0.175, -0.009)** |
| December |  | 0.030 (-0.054, 0.114) | 0.024 (-0.061, 0.109) |
| Year of entry (ref: 2014) |  |  |  |
| 2015 |  | 0.004 (-0.073, 0.080) | 0.003 (-0.081, 0.075) |
| 2016 |  | 0.038 (-0.047, 0.123) | 0.025 (-0.064, 0.115) |
| 2017 |  | 0.001 (-0.076, 0.078) | 0.016 (-0.103, 0.071) |
| 2018 |  | -0.051 (-0.130, 0.028) | -0.075 (-0.173, 0.024) |
| 2019 |  | - | - |
| Most affected joint is hip (ref: knee) |  | **0.095 (0.051, 0.139)** | **0.095 (0.051, 0.138)** |
| BMI (kg/m^2^) |  | **0.025 (0.005, 0.045)** | **0.025 (0.005, 0.045)** |
| Duration of symptoms (months) |  | **0.054 (0.035, 0.073)** | **0.054 (0.035, 0.073)** |
| 40m walk test at baseline |  | **-0.051 (-0.076, -0.025)** | **-0.052 (-0.077, -0.027)** |
| No. of chair stands during 30sec at baseline |  | **-0.029 (-0.051, -0.007)** | **-0.029 (-0.051, -0.007)** |
| No. of painful body areas (0-56) at baseline |  | **0.057 (0.037, 0.077)** | **0.057 (0.037, 0.076)** |
| UCLA - Physical activity (1-10) at baseline |  | 0.004 (-0.017, 0.024) | 0.004 (-0.017, 0.024) |
| KOOS/HOOS QOL (0-100) at baseline |  | **-0.074 (-0.097, -0.050)** | **-0.073 (-0.097, -0.050)** |
| ASES: Other (10-100) at baseline |  | -0.001 (-0.033, 0.031) | -0.001 (-0.033, 0.031) |
| ASES: Pain (10-100) at baseline |  | **-0.132 (-0.161, -0.103)** | **-0.132 (-0.161, -0.103)** |
| EQ-VAS (0-100) at baseline |  | **-0.038 (-0.061, -0.014)** | **-0.038 (-0.061, -0.014)** |
| EQ-5D Health Utility score (-0.757-1) at baseline |  | **-0.082 (-0.108, -0.057)** | **-0.082 (-0.108, -0.056)** |
| SF-12 PCS (0-100) at baseline |  | 0.002 (-0.023, 0.028) | 0.002 (-0.024, 0.027) |
| SF-12 MCS (0-100) at baseline |  | **0.023 (0.000, 0.045)** | **0.023 (0.000, 0.045)** |
| Pain intensity (0-100) at baseline |  | **0.165 (0.144, 0.186)** | **0.165 (0.144, 0.186)** |
| VPC (ICC) | 0.015  (0.006, 0.023) | 0.016  (0.005, 0.025) | 0.016  (0.007, 0.022) |
| AIC | 27569.49 | 26206.81 | 26220.89 |
| BIC | 27591.03 | 26472.55 | 26500.99 |
| NB All predictors standardised and centred  **Adj.b** Adjusted beta coefficient; **AIC** Akaike Information Criterion; **ASES** Arthritis Self-Efficacy Scale; **BIC** Bayesian Information Criterion; **EQ5D** EuroQoL-5 dimensions; **HOOS** Hip Osteoarthritis Outcome Score; **ICC** Intra-class Correlation Coefficient; **IQR** Interquartile range; **KOOS** Knee Osteoarthritis Outcome Score; **MCS** Mental Component Score; **PCS** Physical Component Score; **QOL** Quality of Life; **SD** Standard deviation; **UCLA** University of California, Los Angeles; **VAS** Visual Analogue Score; **VPC** Variance Partition Coefficient; **95%CI** 95 percent confidence interval | | | |

**Table S10.** Random intercept linear regression models for HOOS/KOOS Quality of Life Score (0-100) at 3 months: complete case analysis

|  | Model 0 | Model 1 | Model 2 |
| --- | --- | --- | --- |
|  | ‘Null’ model | Patient-level adjustment model | ‘Full model’ |
|  | N=9,720 | N=9,720 | N=9,720 |
|  |  | Adj.b (95%CI) | Adj.b (95%CI) |
| **Therapist-level variables** |  |  |  |
| Number of patients treated |  |  | -0.005 (-0.032, 0.022) |
| Days since therapist certification |  |  | 0.016 (-0.012, 0.045) |
| **Patient-level variables** |  |  |  |
| Age |  | 0.006 (-0.013, 0.025) | 0.006 (-0.013, 0.025) |
| Female sex |  | **0.086 (0.047, 0.125)** | **0.086 (0.047, 0.125)** |
| Born in Denmark |  | 0.000 (-0.110, 0.110) | 0.000 (-0.110, 0.110) |
| Danish citizen |  | -0.006 (-0.174, 0.163) | -0.005 (-0.173, 0.164) |
| Month of entry (ref: January) |  |  |  |
| February |  | -0.014 (-0.087, 0.059) | -0.012 (-0.085, 0.061) |
| March |  | 0.070 (-0.001, 0.142) | 0.070 (-0.001, 0.142) |
| April |  | 0.060 (-0.010, 0.129) | 0.059 (-0.010, 0.129) |
| May |  | 0.056 (-0.022, 0.133) | 0.055 (-0.022, 0.133) |
| June |  | 0.046 (-0.047, 0.138) | 0.044 (-0.048, 0.137) |
| July |  | 0.027 (-0.071, 0.124) | 0.025 (-0.073, 0.123) |
| August |  | 0.015 (-0.054, 0.085) | 0.011 (-0.059, 0.081) |
| September |  | 0.038 (-0.039, 0.115) | 0.034 (-0.043, 0.111) |
| October |  | 0.057 (-0.015, 0.128) | 0.052 (-0.020, 0.124) |
| November |  | 0.044 (-0.030, 0.118) | 0.040 (-0.034, 0.114) |
| December |  | -0.017 (-0.092, 0.058) | -0.023 (-0.099, 0.053) |
| Year of entry (ref: 2014) |  |  |  |
| 2015 |  | 0.031 (-0.037, 0.100) | 0.025 (-0.045, 0.094) |
| 2016 |  | 0.011 (-0.065, 0.087) | -0.002 (-0.081, 0.078) |
| 2017 |  | -0.007 (-0.076, 0.061) | -0.026 (-0.103, 0.051) |
| 2018 |  | 0.057 (-0.013, 0.127) | 0.029 (-0.057, 0.116) |
| 2019 |  | - | - |
| Most affected joint is hip (ref: knee) |  | 0.011 (-0.028, 0.050) | 0.011 (-0.028, 0.050) |
| BMI (kg/m^2^) |  | **-0.026 (-0.044, -0.008)** | **-0.026 (-0.044, -0.008)** |
| Duration of symptoms (months) |  | **-0.046 (-0.063, -0.029)** | **-0.046 (-0.063, -0.029)** |
| 40m walk test at baseline |  | **0.022 (0.000, 0.045)** | **0.023 (0.001, 0.045)** |
| No. of chair stands during 30sec at baseline |  | **0.024 (0.004, 0.044)** | **0.024 (0.004, 0.043)** |
| No. of painful body areas (0-56) at baseline |  | **-0.029 (-0.047, -0.011)** | **-0.029 (-0.047, -0.011)** |
| UCLA - Physical activity (1-10) at baseline |  | -0.022 (-0.040, -0.004) | -0.022 (-0.040, -0.004) |
| KOOS/HOOS QOL (0-100) at baseline |  | **0.376 (0.355, 0.397)** | **0.376 (0.355, 0.397)** |
| ASES: Other (10-100) at baseline |  | 0.022 (-0.006, 0.051) | 0.022 (-0.006, 0.050) |
| ASES: Pain (10-100) at baseline |  | **0.082 (0.056, 0.108)** | **0.082 (0.056, 0.108)** |
| EQ-VAS (0-100) at baseline |  | **0.027 (0.006, 0.048)** | **0.027 (0.006, 0.048)** |
| EQ-5D Health Utility score (-0.757-1) at baseline |  | **0.056 (0.033, 0.079)** | **0.056 (0.033, 0.079)** |
| SF-12 PCS (0-100) at baseline |  | **0.101 (0.078, 0.124)** | **0.101 (0.078, 0.124)** |
| SF-12 MCS (0-100) at baseline |  | **0.025 (0.005, 0.045)** | **0.025 (0.005, 0.045)** |
| Pain intensity (0-100) at baseline |  | -0.009 (-0.028, 0.010) | -0.009 (-0.028, 0.010) |
| VPC (ICC) | 0.014  (0.006, 0.026) | 0.012  (0.002, 0.022) | 0.012  (0.003, 0.022) |
| AIC | 27572.00 | 24055.61 | 24071.95 |
| BIC | 27593.55 | 24321.34 | 24352.05 |
| NB All predictors standardised and centred  **Adj.b** Adjusted beta coefficient; **AIC** Akaike Information Criterion; **ASES** Arthritis Self-Efficacy Scale; **BIC** Bayesian Information Criterion; **EQ5D** EuroQoL-5 dimensions; **HOOS** Hip Osteoarthritis Outcome Score; **ICC** Intra-class Correlation Coefficient; **IQR** Interquartile range; **KOOS** Knee Osteoarthritis Outcome Score; **MCS** Mental Component Score; **PCS** Physical Component Score; **QOL** Quality of Life; **SD** Standard deviation; **UCLA** University of California, Los Angeles; **VAS** Visual Analogue Score; **VPC** Variance Partition Coefficient; **95%CI** 95 percent confidence interval | | | |

**Table S11.** Random intercept linear regression models for EQ5D Health Utility Score (-0.757-1.000) at 3 months: complete case analysis

|  | Model 0 | Model 1 | Model 2 |
| --- | --- | --- | --- |
|  | ‘Null’ model | Patient-level adjustment model | ‘Full model’ |
|  | N=9,720 | N=9,720 | N=9,720 |
|  |  | Adj.b (95%CI) | Adj.b (95%CI) |
| **Therapist-level variables** |  |  |  |
| Number of patients treated |  |  | -0.014 (-0.040, 0.013) |
| Days since therapist certification |  |  | 0.011 (-0.017, 0.039) |
| **Patient-level variables** |  |  |  |
| Age |  | 0.000 (-0.019, 0.019) | 0.000 (-0.019, 0.020) |
| Female sex |  | **0.140 (0.100, 0.179)** | **0.140 (0.101, 0.180)** |
| Born in Denmark |  | -0.057 (-0.168, 0.054) | -0.057 (-0.168, 0.054) |
| Danish citizen |  | 0.007 (-0.163, 0.177) | 0.008 (-0.162, 0.178) |
| Month of entry (ref: January) |  |  |  |
| February |  | -0.028 (-0.101, 0.046) | -0.027 (-0.100, 0.047) |
| March |  | 0.002 (-0.069, 0.074) | 0.003 (-0.069, 0.075) |
| April |  | 0.002 (-0.068, 0.073) | 0.002 (-0.068, 0.073) |
| May |  | 0.026 (-0.052, 0.104) | 0.026 (-0.052, 0.104) |
| June |  | 0.020 (-0.073, 0.113) | 0.020 (-0.073, 0.114) |
| July |  | 0.059 (-0.040, 0.157) | 0.060 (-0.039, 0.158) |
| August |  | -0.019 (-0.089, 0.051) | -0.021 (-0.091, 0.049) |
| September |  | -0.006 (-0.084, 0.071) | -0.007 (-0.084, 0.071) |
| October |  | 0.036 (-0.036, 0.108) | 0.035 (-0.037, 0.108) |
| November |  | 0.055 (-0.019, 0.129) | 0.055 (-0.019, 0.130) |
| December |  | -0.010 (-0.086, 0.065) | -0.010 (-0.087, 0.066) |
| Year of entry (ref: 2014) |  |  |  |
| 2015 |  | -0.016 (-0.085, 0.053) | -0.017 (-0.087, 0.053) |
| 2016 |  | 0.002 (-0.075, 0.078) | 0.002 (-0.075, 0.078) |
| 2017 |  | -0.032 (-0.101, 0.036) | -0.034 (-0.110, 0.041) |
| 2018 |  | -0.022 (-0.093, 0.048) | -0.027 (-0.111, 0.058) |
| 2019 |  | - | - |
| Most affected joint is hip (ref: knee) |  | **-0.142 (-0.181, -0.102)** | **-0.142 (-0.181, -0.102)** |
| BMI (kg/m^2^) |  | -0.015 (-0.033, 0.004) | -0.015 (-0.033, 0.004) |
| Duration of symptoms (months) |  | **-0.024 (-0.041, -0.007)** | **-0.024 (-0.041, -0.007)** |
| 40m walk test at baseline |  | **0.031 (0.009, 0.054)** | **0.033 (0.010, 0.055)** |
| No. of chair stands during 30sec at baseline |  | **0.022 (0.002, 0.042)** | **0.021 (0.001, 0.041)** |
| No. of painful body areas (0-56) at baseline |  | **-0.040 (-0.058, -0.022)** | **-0.040 (-0.058, -0.022)** |
| UCLA - Physical activity (1-10) at baseline |  | -0.001 (-0.020, 0.017) | -0.001 (-0.020, 0.017) |
| KOOS/HOOS QOL (0-100) at baseline |  | **0.081 (0.059, 0.102)** | **0.080 (0.059, 0.102)** |
| ASES: Other (10-100) at baseline |  | **0.063 (0.034, 0.092)** | **0.063 (0.034, 0.091)** |
| ASES: Pain (10-100) at baseline |  | **0.054 (0.028, 0.080)** | **0.054 (0.028, 0.080)** |
| EQ-VAS (0-100) at baseline |  | **0.043 (0.022, 0.064)** | **0.043 (0.022, 0.064)** |
| EQ-5D Health Utility score (-0.757-1) at baseline |  | **0.298 (0.275, 0.321)** | **0.298 (0.274, 0.321)** |
| SF-12 PCS (0-100) at baseline |  | **0.081 (0.058, 0.104)** | **0.081 (0.058, 0.104)** |
| SF-12 MCS (0-100) at baseline |  | **0.059 (0.039, 0.080)** | **0.059 (0.039, 0.080)** |
| Pain intensity (0-100) at baseline |  | -0.025 (0.044, -0.006) | -0.025 (0.044, -0.006) |
| VPC (ICC) | 0.012  (0.001, 0.022) | 0.008  (0.001, 0.016) | 0.008  (0.000, 0.019) |
| AIC | 27578.86 | 24215.12 | 24231.9 |
| BIC | 27600.41 | 24480.85 | 24511.99 |
| NB All predictors standardised and centred  **Adj.b** Adjusted beta coefficient; **AIC** Akaike Information Criterion; **ASES** Arthritis Self-Efficacy Scale; **BIC** Bayesian Information Criterion; **EQ5D** EuroQoL-5 dimensions; **HOOS** Hip Osteoarthritis Outcome Score; **ICC** Intra-class Correlation Coefficient; **IQR** Interquartile range; **KOOS** Knee Osteoarthritis Outcome Score; **MCS** Mental Component Score; **PCS** Physical Component Score; **QOL** Quality of Life; **SD** Standard deviation; **UCLA** University of California, Los Angeles; **VAS** Visual Analogue Score; **VPC** Variance Partition Coefficient; **95%CI** 95 percent confidence interval | | | |

**Table S12.** Random intercept linear regression models for EQ5D VAS (0-100) at 3 months: complete case analysis

|  | Model 0 | Model 1 | Model 2 |
| --- | --- | --- | --- |
|  | ‘Null’ model | Patient-level adjustment model | ‘Full model’ |
|  | N=9,720 | N=9,720 | N=9,720 |
|  |  | Adj.b (95%CI) | Adj.b (95%CI) |
| **Therapist-level variables** |  |  |  |
| Number of patients treated |  |  | 0.011 (-0.015, 0.038) |
| Days since therapist certification |  |  | 0.010 (-0.018, 0.037) |
| **Patient-level variables** |  |  |  |
| Age |  | 0.004 (-0.015, 0.023) | 0.004 (-0.016, 0.023) |
| Female sex |  | **0.094 (0.054, 0.133)** | **0.093 (0.054, 0.132)** |
| Born in Denmark |  | 0.042 (-0.069, 0.152) | 0.042 (-0.068, 0.153) |
| Danish citizen |  | 0.060 (-0.109, 0.230) | 0.061 (-0.109, 0.230) |
| Month of entry (ref: January) |  |  |  |
| February |  | 0.022 (-0.052, 0.095) | 0.023 (-0.050, 0.096) |
| March |  | 0.049 (-0.023, 0.120) | 0.048 (-0.024, 0.119) |
| April |  | 0.020 (-0.050, 0.090) | 0.020 (-0.050, 0.090) |
| May |  | 0.050 (-0.028, 0.128) | 0.049 (-0.029, 0.127) |
| June |  | -0.019 (-0.112, 0.074) | -0.022 (-0.115, 0.071) |
| July |  | -0.012 (-0.089, 0.066) | -0.015 (-0.113, 0.084) |
| August |  | -0.005 (-0.075, 0.065) | -0.009 (-0.079, 0.061) |
| September |  | -0.012 (-0.089, 0.066) | -0.017 (-0.095, 0.061) |
| October |  | -0.024 (-0.095, 0.048) | -0.029 (-0.101, 0.043) |
| November |  | 0.057 (-0.017, 0.131) | 0.050 (-0.024, 0.125) |
| December |  | 0.038 (-0.038, 0.114) | 0.030 (-0.046, 0.106) |
| Year of entry (ref: 2014) |  |  |  |
| 2015 |  | -0.033 (-0.101, 0.036) | -0.042 (-0.111, 0.028) |
| 2016 |  | -0.061 (-0.137, 0.015) | -0.078 (-0.158, 0.001) |
| 2017 |  | **-0.087 (-0.156, -0.019)** | **-0.111 (-0.187, -0.036)** |
| 2018 |  | **-0.075 (-0.145, -0.005)** | **-0.110 (-0.195, -0.026)** |
| 2019 |  | - | - |
| Most affected joint is hip (ref: knee) |  | **-0.096 (-0.135, -0.057)** | **-0.096 (-0.136, -0.057)** |
| BMI (kg/m^2^) |  | **-0.028 (-0.046, -0.010)** | **-0.028 (-0.046, -0.010)** |
| Duration of symptoms (months) |  | **-0.024 (-0.041, -0.007)** | **-0.025 (-0.042, -0.008)** |
| 40m walk test at baseline |  | 0.019 (-0.003, 0.042) | 0.019 (-0.004, 0.041) |
| No. of chair stands during 30sec at baseline |  | **0.030 (0.010, 0.049)** | **0.029 (0.009, 0.049)** |
| No. of painful body areas (0-56) at baseline |  | **-0.056 (-0.074, -0.039)** | **-0.056 (-0.074, -0.038)** |
| UCLA - Physical activity (1-10) at baseline |  | **0.018 (0.000, 0.036)** | **0.018 (0.000, 0.037)** |
| KOOS/HOOS QOL (0-100) at baseline |  | **0.038 (0.017, 0.059)** | **0.038 (0.017, 0.059)** |
| ASES: Other (10-100) at baseline |  | **0.095 (0.067, 0.124)** | **0.095 (0.067, 0.124)** |
| ASES: Pain (10-100) at baseline |  | **0.051 (0.025, 0.077)** | **0.051 (0.025, 0.077)** |
| EQ-VAS (0-100) at baseline |  | **0.291 (0.270, 0.312)** | **0.291 (0.270, 0.312)** |
| EQ-5D Health Utility score (-0.757-1) at baseline |  | **0.026 (0.003, 0.049)** | **0.026 (0.003, 0.049)** |
| SF-12 PCS (0-100) at baseline |  | **0.114 (0.091, 0.137)** | **0.113 (0.090, 0.136)** |
| SF-12 MCS (0-100) at baseline |  | **0.070 (0.050, 0.090)** | **0.070 (0.049, 0.090)** |
| Pain intensity (0-100) at baseline |  | -0.009 (-0.027, 0.010) | -0.009 (-0.028, 0.010) |
| VPC (ICC) | 0.015  (0.001, 0.026) | 0.008  (0.001, 0.015) | 0.008  (0.002, 0.018) |
| AIC | 27559.49 | 24161.87 | 24176.97 |
| BIC | 27581.03 | 24427.6 | 24457.07 |
| NB All predictors standardised and centred  **Adj.b** Adjusted beta coefficient; **AIC** Akaike Information Criterion; **ASES** Arthritis Self-Efficacy Scale; **BIC** Bayesian Information Criterion; **EQ5D** EuroQoL-5 dimensions; **HOOS** Hip Osteoarthritis Outcome Score; **ICC** Intra-class Correlation Coefficient; **IQR** Interquartile range; **KOOS** Knee Osteoarthritis Outcome Score; **MCS** Mental Component Score; **PCS** Physical Component Score; **QOL** Quality of Life; **SD** Standard deviation; **UCLA** University of California, Los Angeles; **VAS** Visual Analogue Score; **VPC** Variance Partition Coefficient; **95%CI** 95 percent confidence interval | | | |

**Table S13.** Random intercept logistic regression models for ≥30% pain reduction at 3 months: complete case analysis

|  | Model 0 | Model 1 | Model 2 |
| --- | --- | --- | --- |
|  | ‘Null’ model | Patient-level adjustment model | ‘Full model’ |
|  | N=9,720 | N=9,720 | N=9,720 |
|  |  | aOR (95%CI) | aOR (95%CI) |
| **Therapist-level variables** |  |  |  |
| Number of patients treated |  |  | 0.96 (0.90, 1.03) |
| Days since therapist certification |  |  | 0.99 (0.92, 1.07) |
| **Patient-level variables** |  |  |  |
| Age |  | **0.94 (0.90, 0.99)** | **0.94 (0.90, 0.99)** |
| Female sex |  | **1.25 (1.13, 1.38)** | **1.25 (1.13, 1.38)** |
| Born in Denmark |  | 1.07 (0.81, 1.42) | 1.07 (0.81, 1.42) |
| Danish citizen |  | 1.18 (0.76, 1.82) | 1.18 (0.76, 1.82) |
| Month of enrollment on GLA:D (ref: January) |  |  |  |
| February |  | 0.94 (0.78, 1.13) | 0.94 (0.78, 1.13) |
| March |  | **1.27 (1.06, 1.53)** | **1.28 (1.06, 1.53)** |
| April |  | 1.14 (0.96, 1.36) | 1.14 (0.96, 1.37) |
| May |  | 1.17 (0.96, 1.43) | 1.17 (0.96, 1.43) |
| June |  | 1.10 (0.87, 1.40) | 1.11 (0.87, 1.40) |
| July |  | 1.19 (0.93, 1.53) | 1.20 (0.94, 1.55) |
| August |  | 1.10 (0.92, 1.31) | 1.11 (0.93, 1.32) |
| September |  | 1.13 (0.93, 1.37) | 1.14 (0.94, 1.39) |
| October |  | 1.12 (0.93, 1.34) | 1.13 (0.94, 1.36) |
| November |  | **1.23 (1.02, 1.49)** | **1.25 (1.03, 1.51)** |
| December |  | 1.00 (0.82, 1.21) | 1.01 (0.83, 1.23) |
| Year of enrollment on GLA:D (ref: 2014) |  |  |  |
| 2015 |  | 1.02 (0.86, 1.21) | 1.04 (0.87, 1.24) |
| 2016 |  | 0.96 (0.79, 1.17) | 1.00 (0.81, 1.22) |
| 2017 |  | 1.03 (0.86, 1.22) | 1.08 (0.89, 1.31) |
| 2018 |  | **1.20 (1.00, 1.43)** | **1.28 (1.03, 1.60)** |
| 2019 |  | - | - |
| Most affected joint is hip (ref: knee) |  | **0.86 (0.78, 0.95)** | **0.86 (0.78, 0.95)** |
| BMI (kg/m^2^) |  | 0.96 (0.92, 1.01) | 0.96 (0.92, 1.01) |
| Duration of symptoms (months) |  | **0.91 (0.87, 0.95)** | **0.91 (0.87, 0.95)** |
| 40m walk test at baseline |  | **1.12 (1.05, 1.18)** | **1.12 (1.06, 1.19)** |
| No. of chair stands during 30sec at baseline |  | 1.05 (0.99, 1.10) | 1.05 (1.00, 1.10) |
| No. of painful body areas (0-56) at baseline |  | **0.89 (0.85, 0.93)** | **0.89 (0.85, 0.93)** |
| UCLA - Physical activity (1-10) at baseline |  | 1.02 (0.98, 1.07) | 1.02 (0.98, 1.07) |
| KOOS/HOOS QOL (0-100) at baseline |  | **1.15 (1.09, 1.22)** | **1.15 (1.09, 1.22)** |
| ASES: Other (10-100) at baseline |  | 0.98 (0.91, 1.05) | 0.98 (0.91, 1.05) |
| ASES: Pain (10-100) at baseline |  | **1.34 (1.25, 1.43)** | **1.34 (1.25, 1.43)** |
| EQ-VAS (0-100) at baseline |  | **1.07 (1.02, 1.13)** | **1.07 (1.02, 1.13)** |
| EQ-5D Health Utility score (-0.757-1) at baseline |  | **1.14 (1.08, 1.21)** | **1.14 (1.08, 1.21)** |
| SF-12 PCS (0-100) at baseline |  | 1.01 (0.96, 1.08) | 1.01 (0.96, 1.08) |
| SF-12 MCS (0-100) at baseline |  | 0.97 (0.92, 1.02) | 0.97 (0.92, 1.02) |
| Pain intensity (0-100) at baseline |  | **1.43 (1.36, 1.50)** | **1.43 (1.36, 1.50)** |
| VPC (ICC) | 0.014  (0.005, 0.026) | 0.015  (0.005, 0.025) | 0.015  (0.005, 0.024) |
| AIC | 13423.96 | 12774.19 | 12776.08 |
| BIC | 13438.32 | 13032.74 | 13048.99 |
| AUC | 0.620 | 0.672 | 0.672 |
| NB All predictors standardised and centred  **AIC** Akaike Information Criterion; **ASES** Arthritis Self-Efficacy Scale; **BIC** Bayesian Information Criterion; **EQ5D** EuroQoL-5 dimensions; **HOOS** Hip Osteoarthritis Outcome Score; **ICC** Intra-class Correlation Coefficient; **IQR** Interquartile range; **KOOS** Knee Osteoarthritis Outcome Score; **MCS** Mental Component Score; **aOR** adjusted Odds Ratio; **PCS** Physical Component Score; **QOL** Quality of Life; **SD** Standard deviation; **UCLA** University of California, Los Angeles; **VAS** Visual Analogue Score; **VPC** Variance Partition Coefficient; **95%CI** 95 percent confidence interval | | | |

**Figure S2.** Funnel plot of therapist-specific case-mix-adjusted pain relief50 outcome at 3 months: multiply imputed data

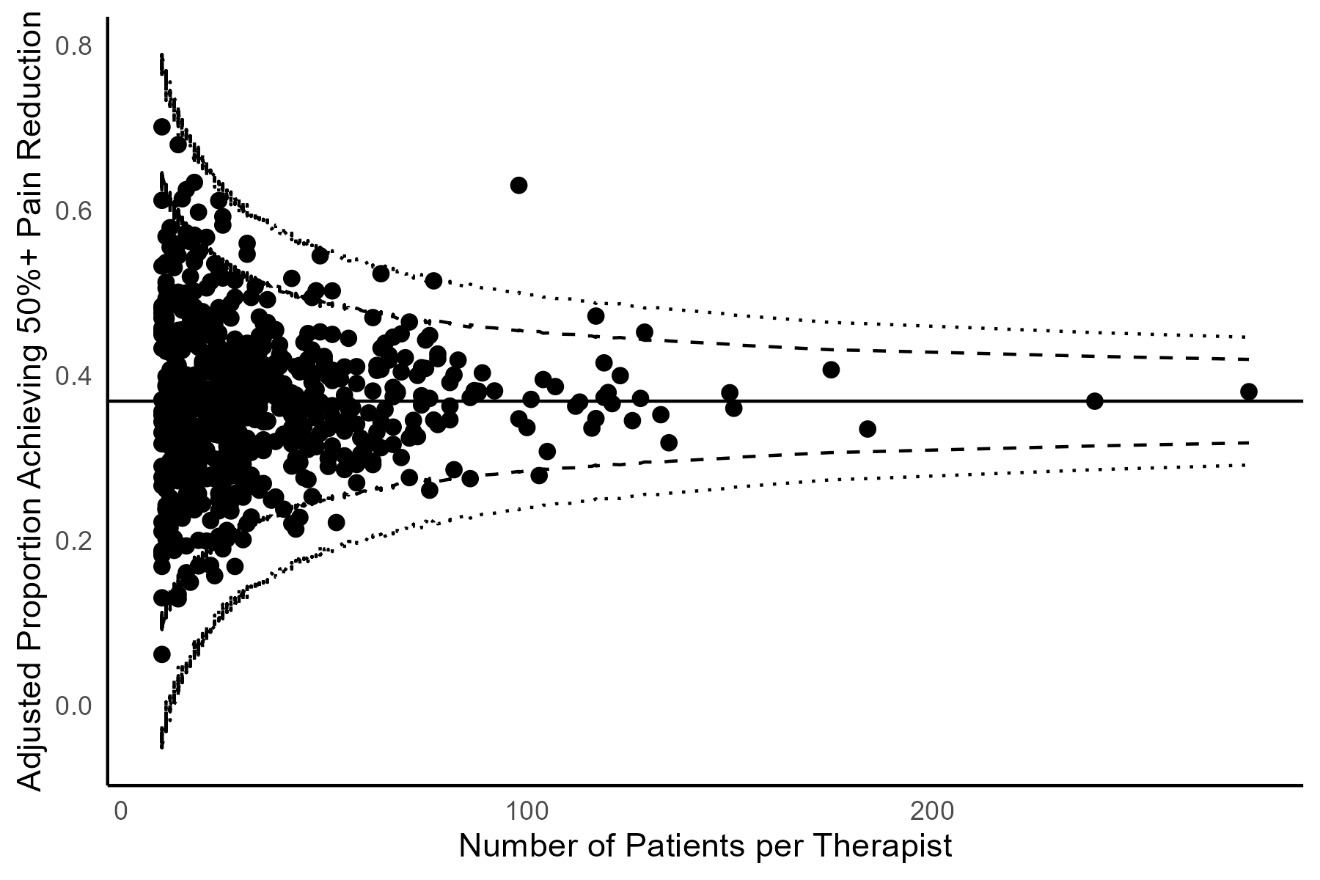

**Figure S3.** Funnel plot of therapist-specific case-mix-adjusted pain intensity (0-100) outcome at 3 months: multiply imputed data

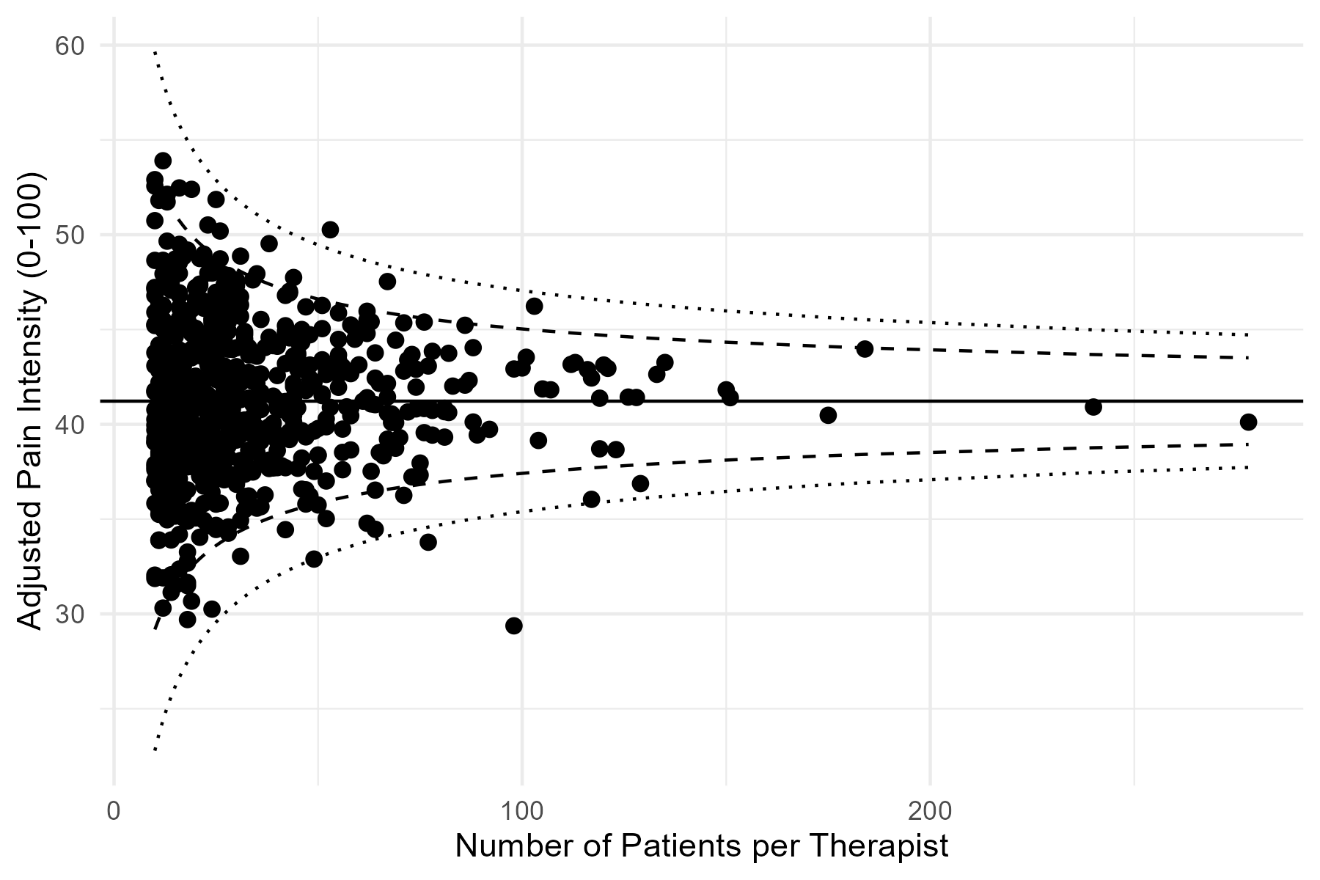

**Figure S4.** Funnel plot of therapist-specific case-mix-adjusted HOOS/KOOS QOL score (0-100) outcome at 3 months: multiply imputed data

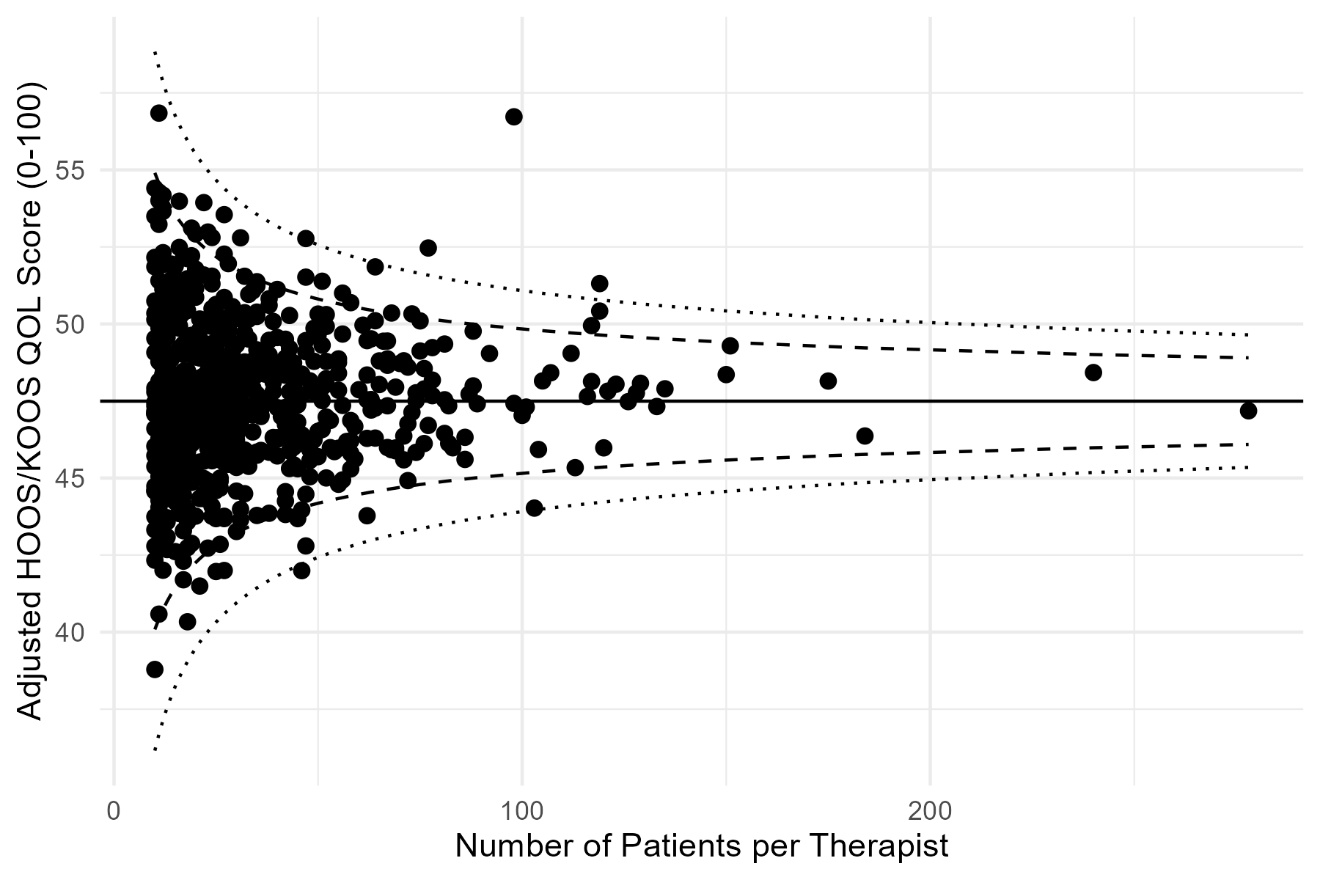

**Figure S5.** Funnel plot of therapist-specific case-mix-adjusted EQ5D Health Utility Score (-0.757-1.000) at 3 months: multiply imputed data

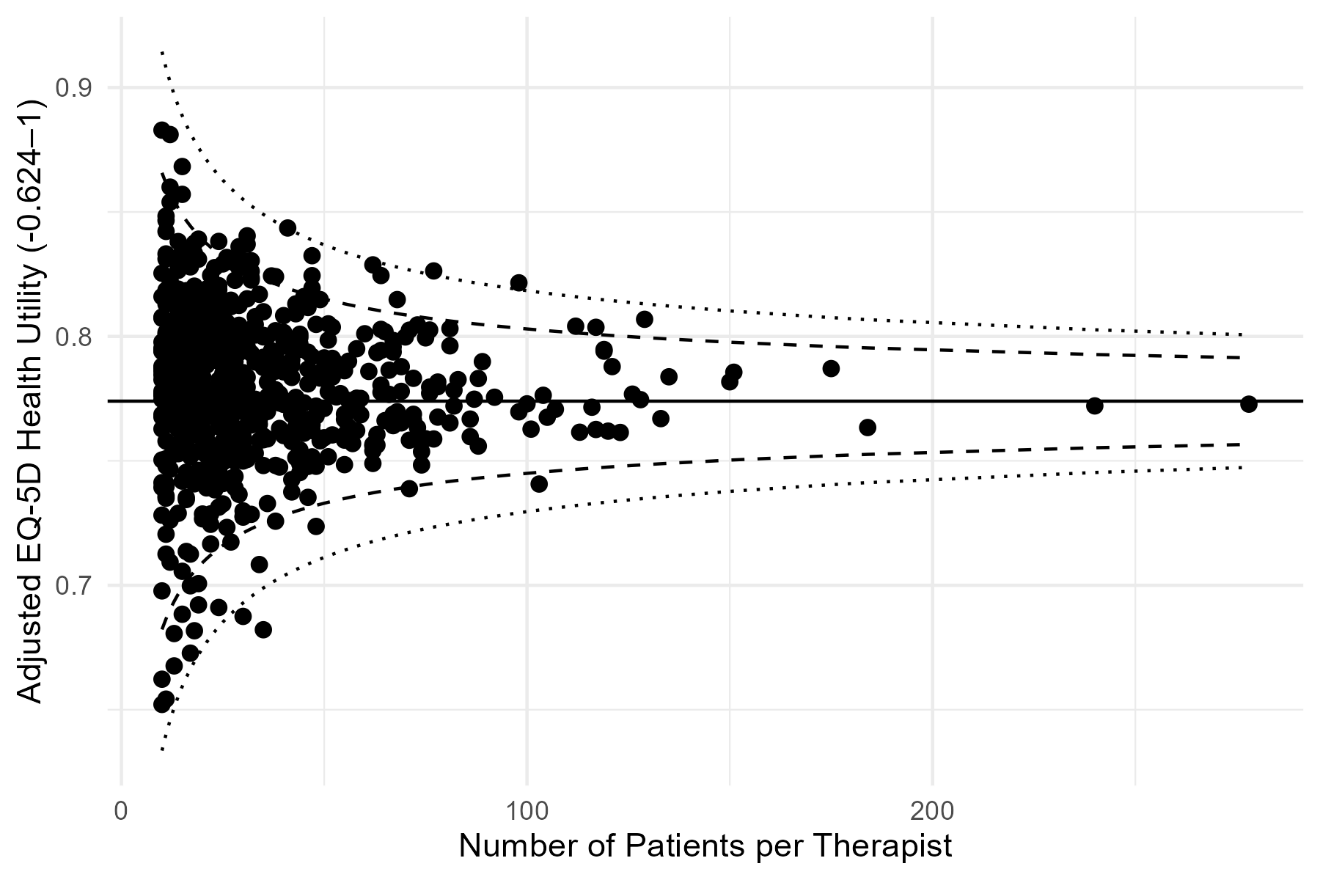

**Figure S6.** Funnel plot of therapist-specific case-mix-adjusted EQ5D VAS (0-100) at 3 months: multiply imputed data

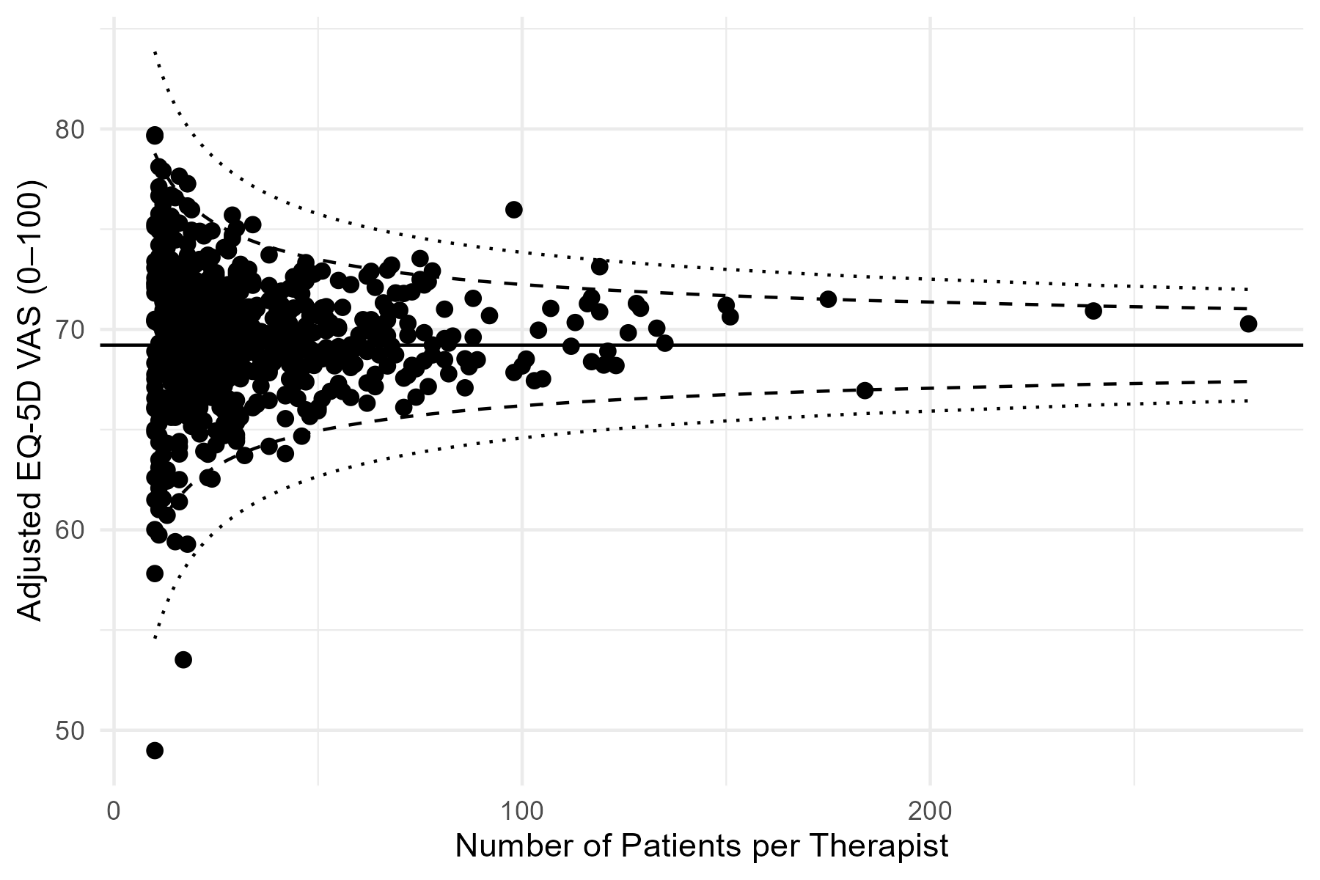

**Figure S7.** Funnel plot of case-mix-adjusted pain relief50 outcome at 3 months: complete case analysis

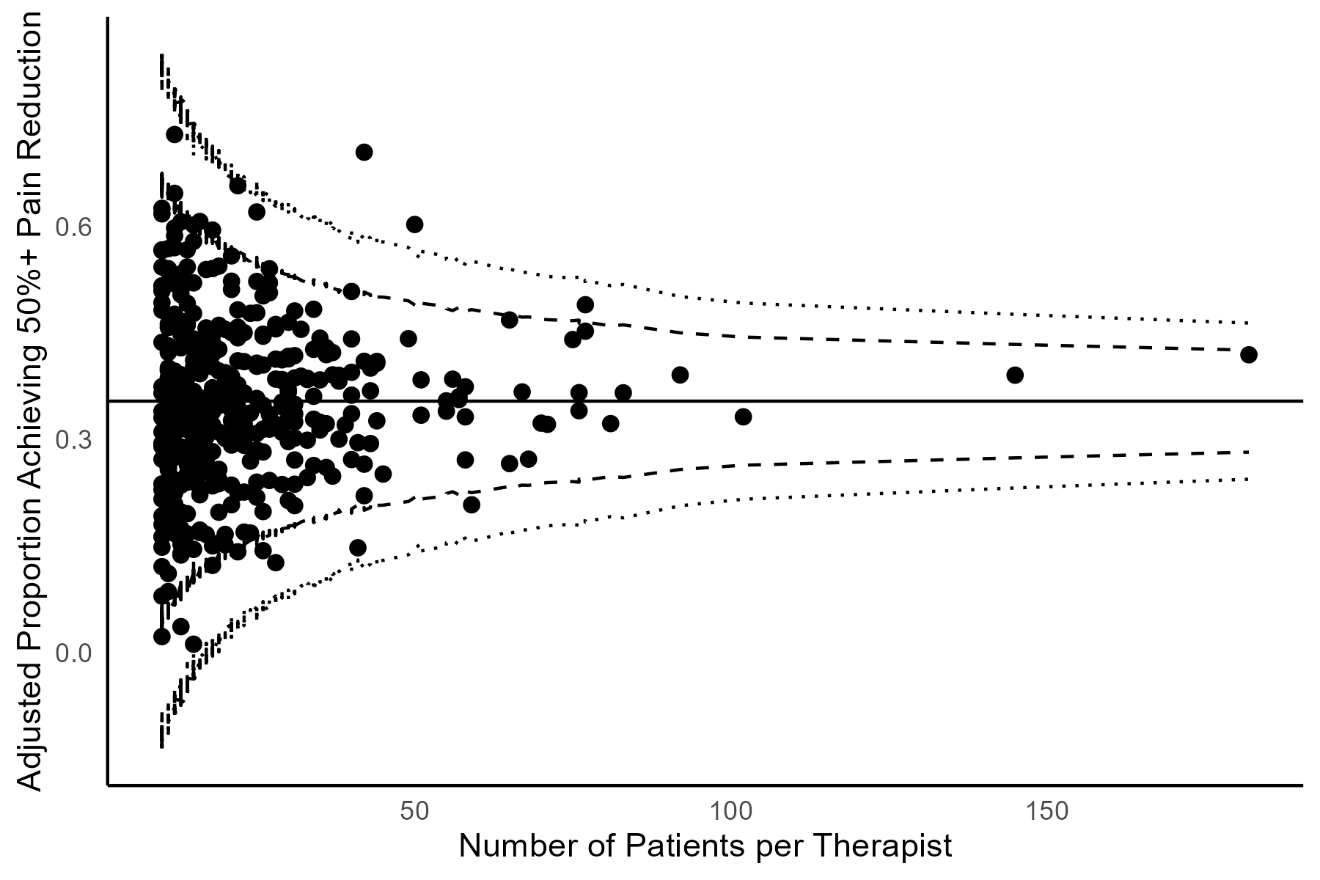

**Figure S8.** Funnel plot of therapist-specific case-mix-adjusted pain intensity (0-100) outcome at 3 months: complete case analysis

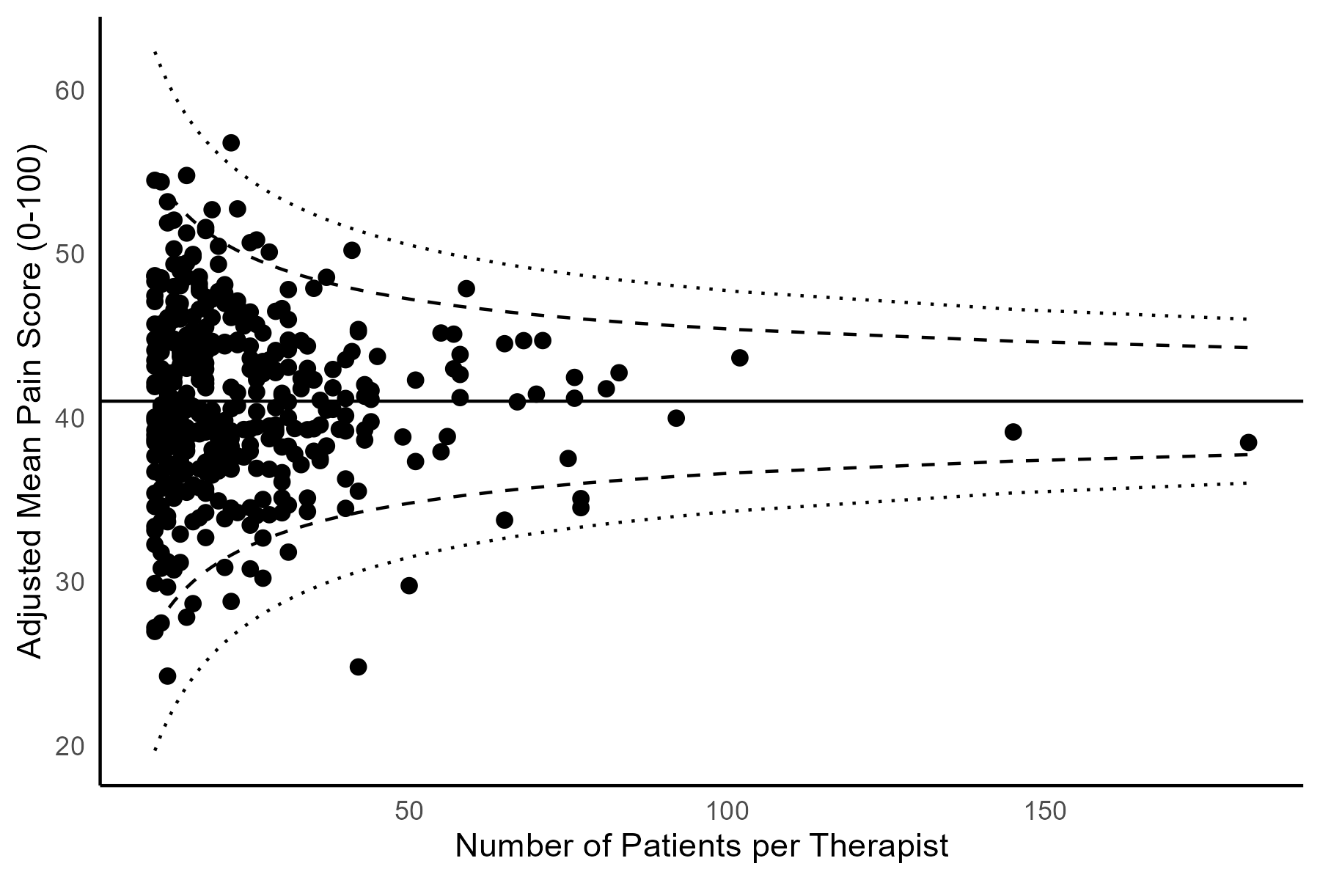

**Figure S9.** Funnel plot of therapist-specific case-mix-adjusted HOOS/KOOS score (0-100) outcome at 3 months: complete case analysis

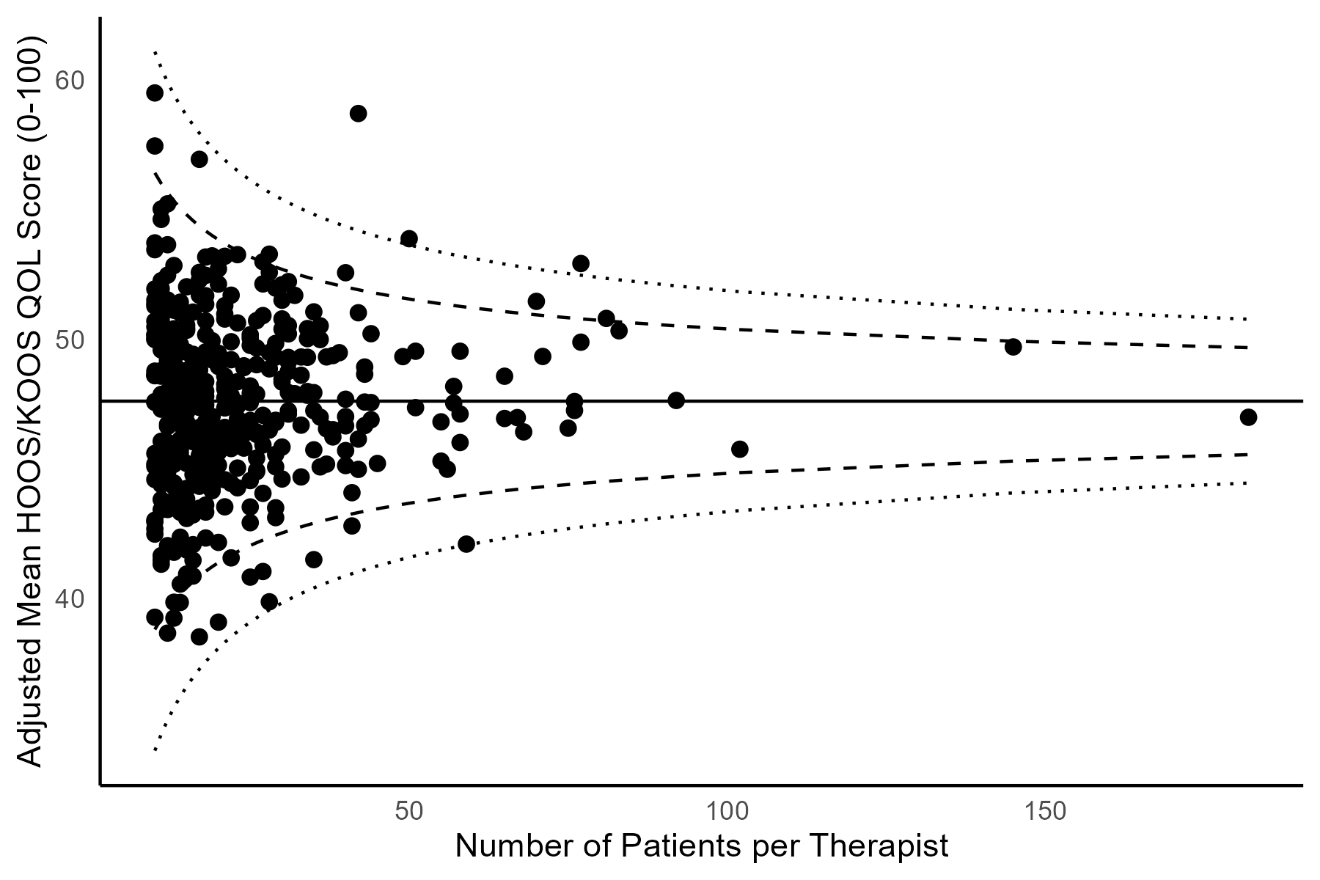

**Figure S10.** Funnel plot of therapist-specific case-mix-adjusted EQ5D Health Utility Score (-0.757-1.000) at 3 months: complete case analysis

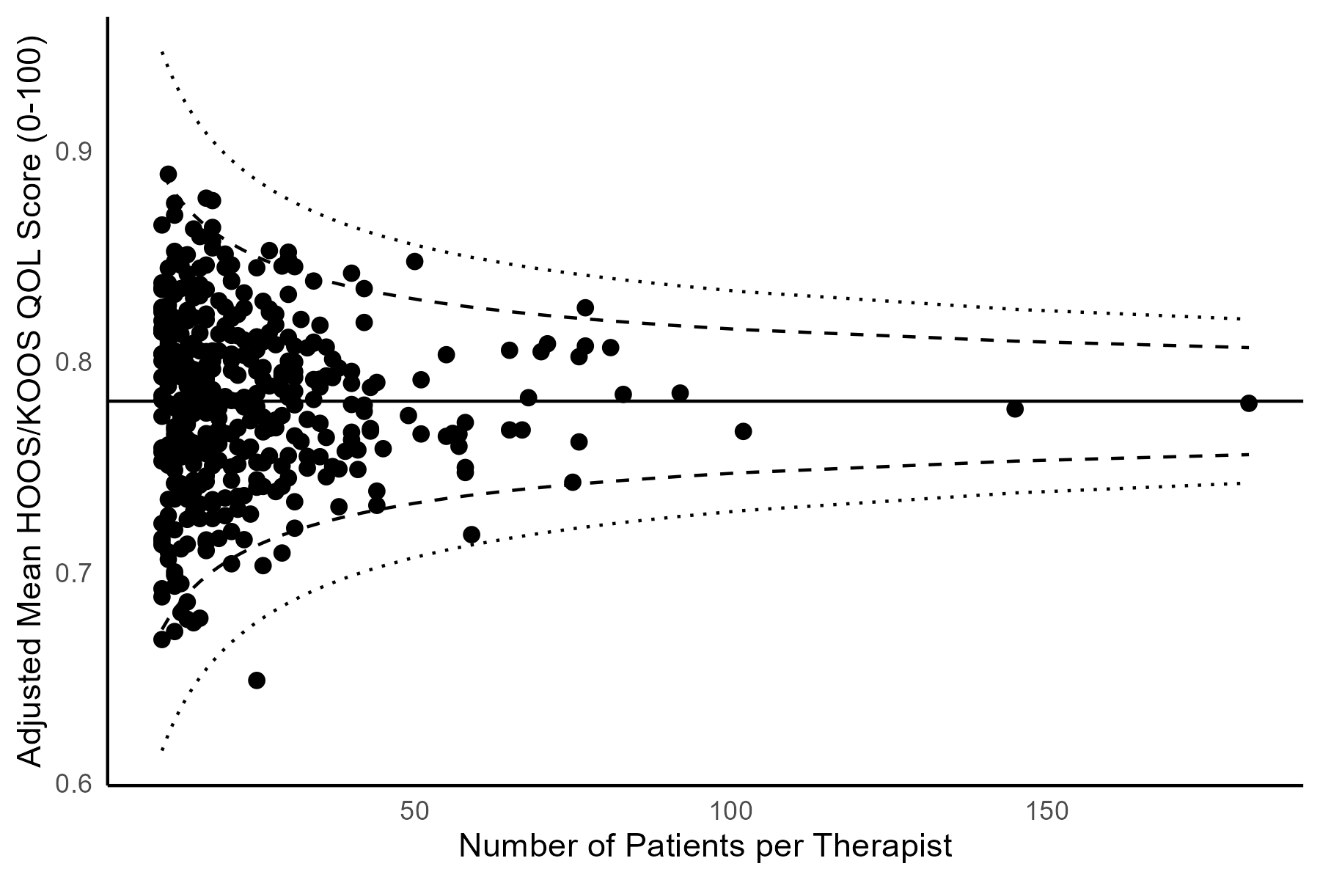

**Figure S11.** Funnel plot of therapist-specific case-mix-adjusted EQ5D VAS (0-100) at 3 months: complete case analysis

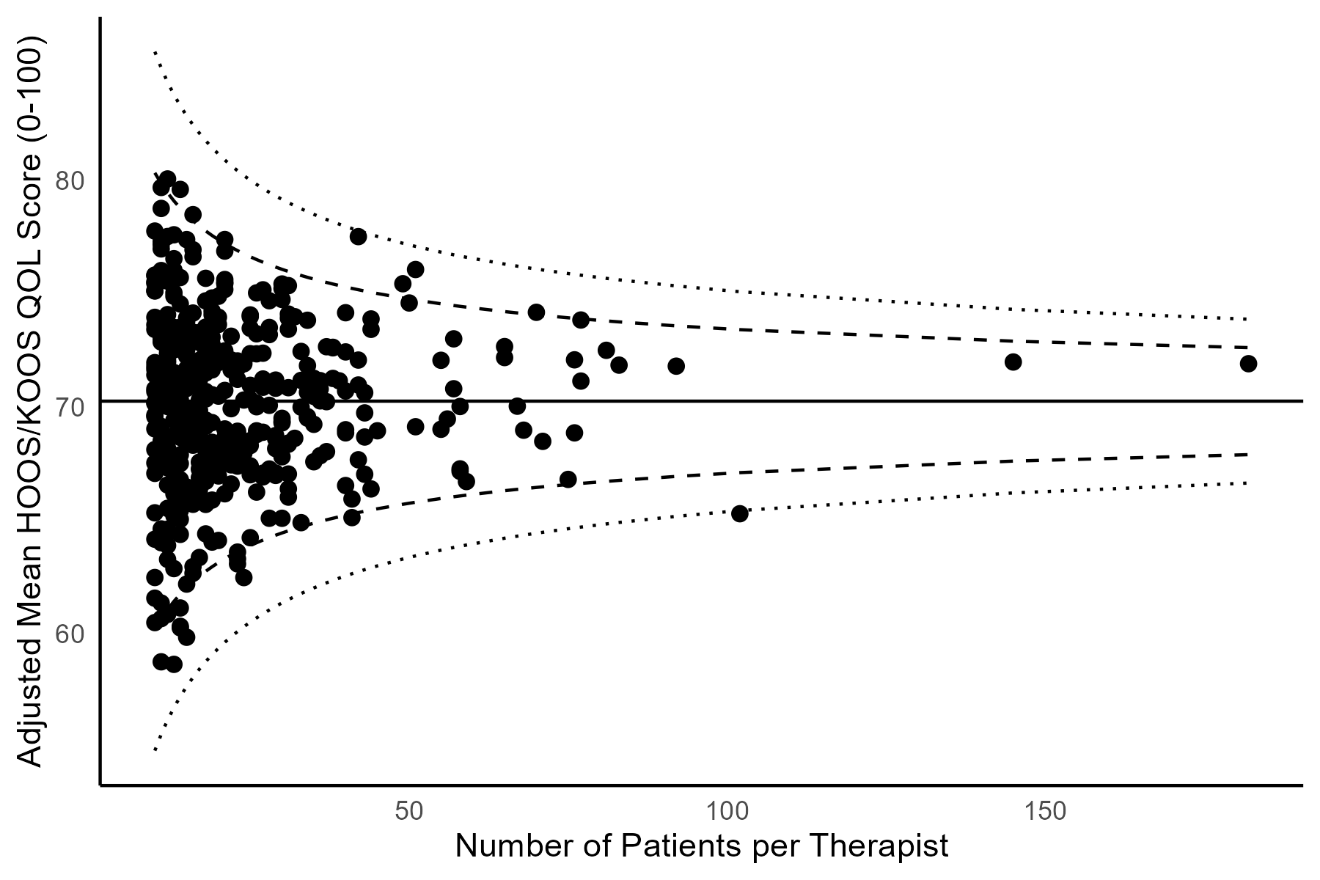

**Figure S12.** Funnel plot of therapist-specific case-mix-adjusted pain relief30 outcome at 3 months: complete case analysis

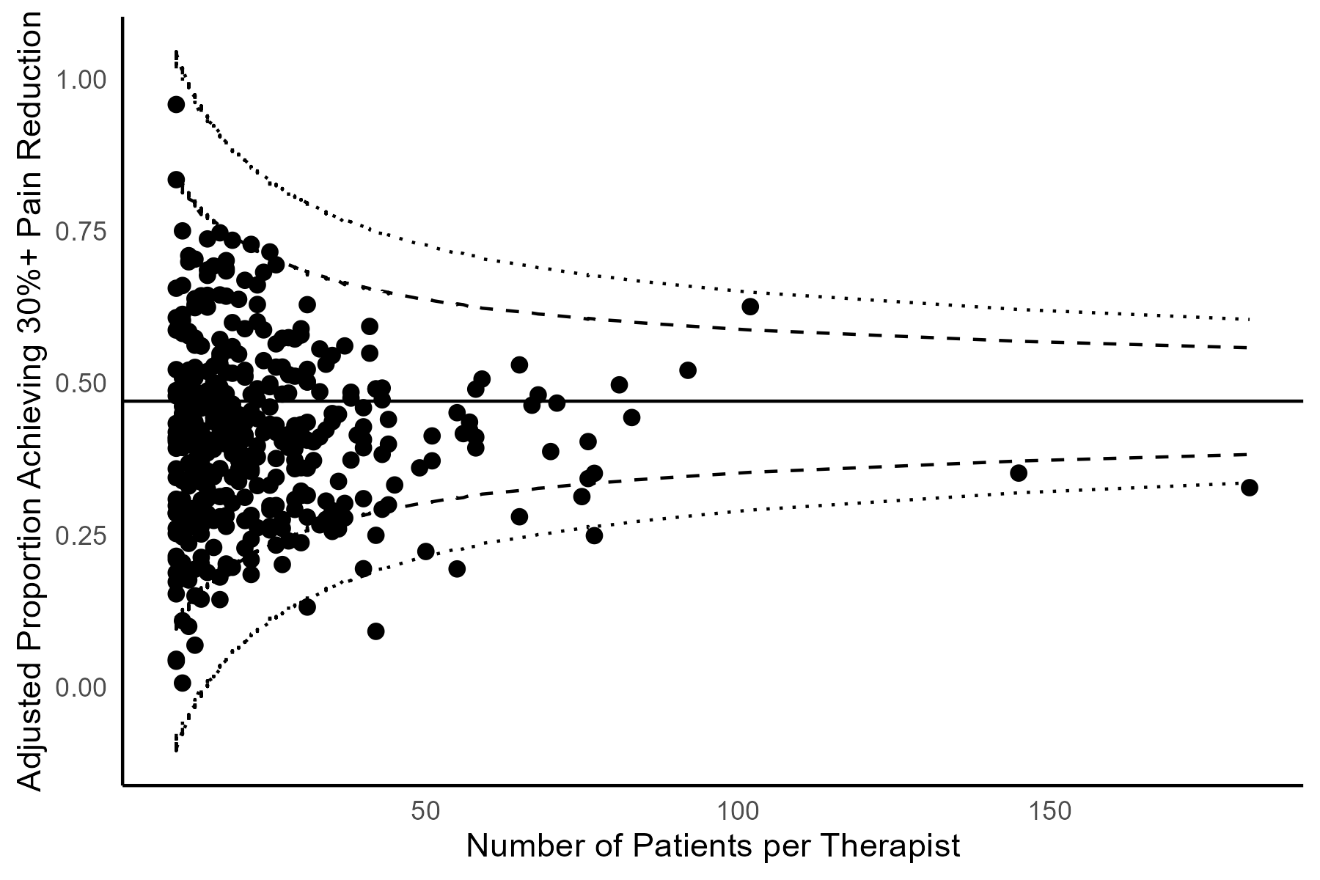

**Figure S13**. Summary of ICC estimates of ‘therapist effects’ for primary and secondary outcomes, multiply imputed data and complete case analysis
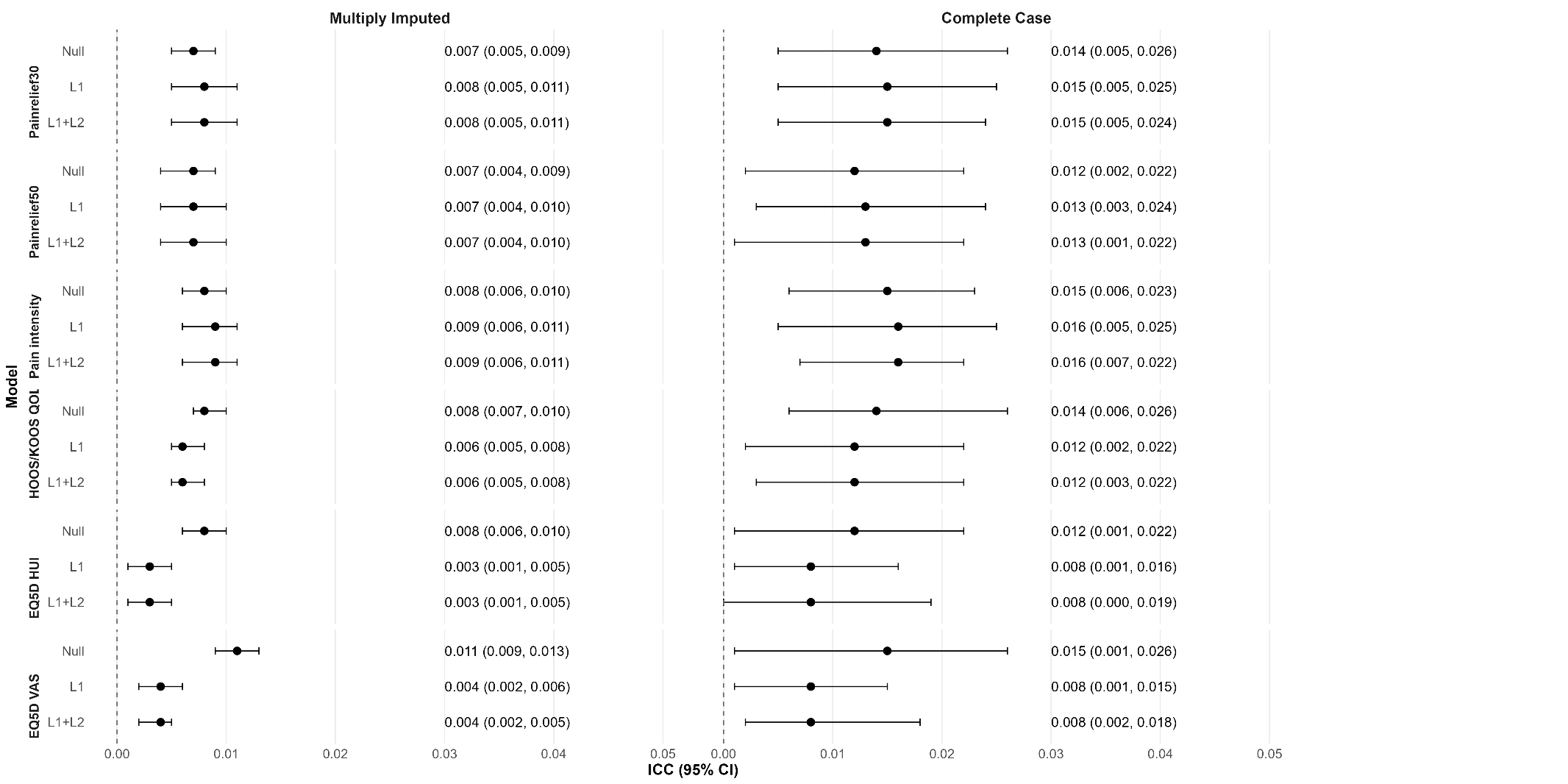
